# Mapping chromatin interactions at melanoma susceptibility loci and cell-type specific dataset integration uncovers distant gene targets of *cis*-regulation

**DOI:** 10.1101/2024.11.14.24317204

**Authors:** Rohit Thakur, Mai Xu, Hayley Sowards, Joshuah Yon, Lea Jessop, Timothy Myers, Tongwu Zhang, Raj Chari, Erping Long, Thomas Rehling, Rebecca Hennessey, Karen Funderburk, Jinhu Yin, Mitchell J. Machiela, Matthew E. Johnson, Andrew D. Wells, Alessandra Chesi, Struan F.A. Grant, Mark M. Iles, Maria Teresa Landi, Matthew H. Law, Melanoma Meta-Analysis Consortium, Jiyeon Choi, Kevin M. Brown

**Author notes:** Contributed equally (Rohit Thakur, Mai Xu).

## Abstract

Genome-wide association studies (GWAS) of melanoma risk have identified 68 independent signals at 54 loci. For most loci, specific functional variants and their respective target genes remain to be established. Capture-HiC is an assay that links fine-mapped risk variants to candidate target genes by comprehensively mapping cell-type specific chromatin interactions. We performed a melanoma GWAS region-focused capture-HiC assay in human primary melanocytes to identify physical interactions between fine-mapped risk variants and potential causal melanoma susceptibility genes. Overall, chromatin interaction data alone nominated potential causal genes for 61 of the 68 melanoma risk signals, identifying many candidates beyond those reported by previous studies. We further integrated these data with cell-type specific epigenomic (chromatin state, accessibility), gene expression (eQTL/TWAS), DNA methylation (meQTL/MWAS), and massively parallel reporter assay (MPRA) data to prioritize potentially *cis*-regulatory variants and their respective candidate gene targets. From the set of fine-mapped variants across these loci, we identified 140 prioritized candidate causal variants linked to 195 candidate genes at 42 risk signals. In addition, we developed an integrative scoring system to facilitate candidate gene prioritization, integrating melanocyte and melanoma datasets. Notably, at several GWAS risk signals we observed long-range chromatin connections (500 kb to >1 Mb) with distant candidate target genes. We validated several such *cis*-regulatory interactions using CRISPR inhibition, providing evidence for known cancer driver genes *MDM4* and *CBL*, as well as the SRY-box transcription factor *SOX4*, as likely melanoma risk genes.

## INTRODUCTION

Melanoma is the deadliest form of skin cancer and originates from melanocytes. Multiple genome-wide association studies (GWAS) of melanoma risk have been conducted^1-9^, with the most recent meta-analysis of 36,760 melanoma cases identifying 54 loci and 68 independent signals^10^. Despite this success, significant challenges lie in pinpointing the functional variants and causal genes at most of these GWAS risk loci^11-13^. Most loci associated with complex traits, including those for melanoma risk, do not harbor risk-associated protein-coding variants^11-13^. Instead, these loci may potentially function through genetic variants located in *cis*-regulatory regions such as enhancers and gene promoters, influencing target gene expression in an allele-specific manner^11-13^. Consistent with this, genetic variants associated with complex traits are often found to be enriched at genomic regions annotated as regulatory elements^14-18^. For many loci, the lead reported variant is not necessarily the functional variant, as each locus may harbor multiple risk-associated variants that are in linkage-disequilibrium (LD) with the unknown causal variant or variants^12^. This often makes it difficult to statistically distinguish the causal risk variant(s) from LD passengers. Furthermore, given that enhancers may function over long distances, the nearest gene to a GWAS risk signal is not necessarily the target gene and therefore, genes other than the nearest gene in the region must be considered plausible targets^11,18,19^.

One commonly used post-GWAS approach for identifying target genes is colocalization of the GWAS signal with those from quantitative trait locus (QTL) datasets generated from disease- or trait-relevant cell types or tissues^20-22^. Colocalization of melanoma GWAS with multiple QTL types derived from expression and methylation data from human primary melanocytes identified at least one colocalizing QTL for less than half (39%) of melanoma GWAS loci^23,24^. While QTLs derived from expression data nominate specific gene candidates for a given locus, meQTLs do not necessarily directly implicate specific genes. Long and colleagues compiled available QTL data with a custom massively parallel reporter assay (MPRA) of fine-mapped melanoma-associated variants and still only linked MPRA-positive variants to genes for roughly 50% of loci^25^. Given many GWAS risk variants have relatively small effects on disease risk, this lack of colocalizing QTLs could be explained by limited statistical power in small QTL studies^11,12,26-28^. Alternatively, there is growing evidence that many variants associated with complex traits may function in a context- and/or state-specific manner which may be missed when using QTL data from cells in a steady-state or MPRA assays in specific cell systems^22,29-31^.

Enhancer elements regulate gene expression via physical interactions with promoter elements and can thus regulate expression of distant genes via long-range three-dimensional chromatin interaction. Methods to characterize chromatin conformation, including HiC-based methods^32-37^ have emerged as powerful approaches to map such interactions at GWAS risk loci^34,35,38-43^. One of these methods, capture-HiC^43-46^, utilizes capture baits targeting regions of interest, often gene promoters or GWAS signals. To date, targeted cell-type specific chromatin interactions have not been evaluated across all genome-wide significant melanoma risk loci, but the utility of this approach has been demonstrated in establishing *AHR* as a functionally-validated ultraviolet B (UVB)-responsive melanoma susceptibility gene^47^. In this study, we performed a GWAS region-specific capture-HiC assay, baiting the entire regions of association for the 68 melanoma GWAS risk signals (locus and signal numbering is listed in **Table S1**) to comprehensively map cell-type specific chromatin interactions between fine-mapped risk variants and potential target genes in human primary melanocytes. We integrated capture-HiC data with fine-mapping, as well as cell-type specific epigenomic (chromatin state, accessibility), gene expression (eQTL/TWAS), DNA methylation (meQTL/MWAS), and high-throughput screening (massively parallel reporter assays; MPRA) data to prioritize variants and their respective candidate gene target(s) for *cis*-regulation (**Figure 1A**).

**Figure 1.**
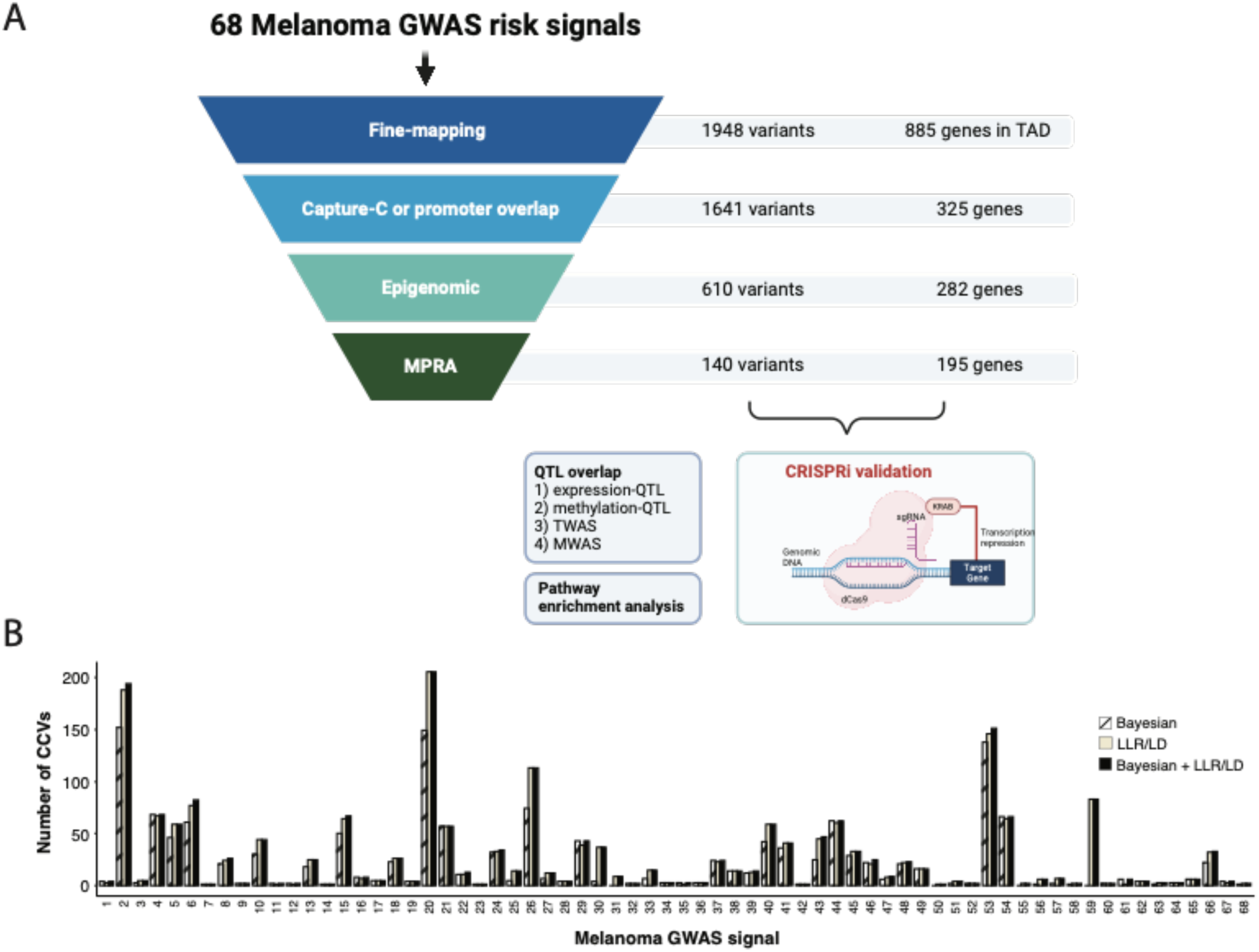
Schematic of data integration of capture-HiC data with orthogonal data to prioritize candidate causal variants and genes. (A) Schematic summary of this study utilizing an integrative analysis approach to identify candidate causal variants (CCVs) and target candidate genes at the 68 melanoma GWAS risk signals. We performed GWAS region-specific capture-HiC assay, baiting the entire region of association for the 68 melanoma GWAS risk signals to comprehensively map chromatin interactions. Subsequently we utilized this dataset to link fine-mapped risk variants to candidate target genes. We integrated fine-mapping with observed chromatin interactions, further overlaying cell-type specific epigenomic (chromatin state, accessibility) and high-throughput reporter assay screening (massively parallel reporter assays, MPRA) datasets to prioritize likely functional variants and respective candidate gene target(s) for *cis*-regulation. Finally, we validated candidate genes nominated at multiple loci via CRISPR inhibition system. (B) Summary of fine-mapped credible causal variants (CCVs) using Bayesian, LLR/LD, or both criteria at the 68 melanoma GWAS signals (a key to numbered loci is provided in **Table S1**). The black bar shows the union of fine-mapped variants identified by Bayesian and LLR/LD approaches.

Our approach nominated potential causal genes for the vast majority of melanoma risk signals, identifying many plausible candidates beyond those reported by previous studies^10,23,24,48^. Notably, we identify multiple candidate genes previously identified as somatically altered in melanoma and pan-cancer tumor analyses.

## METHODS

### Fine-mapping of melanoma risk signals

We performed statistical fine-mapping for the 54 GWAS loci (harboring 68 independent genome-wide significant signals) described by Landi and colleagues^10^ (**Table S1**) using an inclusive strategy, selecting any variant identified by any one of multiple approaches. Firstly, we used a combination of log-likelihood ratio (LLR) and linkage disequilibrium (LD) based cut-offs, similar to that performed for a recent melanoma GWAS massively parallel reporter assay (MPRA) study^25^. The melanoma GWAS summary data was from the fixed-effect inverse-variance weighted meta-analysis of the full set of confirmed and self-reported melanoma cases and controls as previously described^10^.

Specifically, we selected variants that met any one of the following criteria:

a. Variants with log likelihood ratio (LLR) <1:100 relative to the lead variant for the primary signal of each GWAS locus.
b. Variants that were not genotyped or successfully imputed in the GWAS (including insertion/deletion variants not assessed in the Haplotype Reference Consortium imputation panel) that had LD r^2^ > 0.8 (1000 Genomes Project, Phase 3, Version 5, EUR population)^49^ with the primary lead variant. These variants were identified using the LDlinkR package^50-52^.
c. For secondary risk signals at a locus that were identified through conditional analysis within 1 Mb of a primary lead SNP^10^ (irrespective of LLR), we relied on LD-based fine-mapping, selecting all variants with LD r^2^ > 0.8 (based on 1000 Genomes Project, Phase 3, Version 5, EUR population)^49^ to the leading variant at the independent risk signal.

These LLR and LD based data were used for the capture-HiC bait design as well as for identifying credible causal variants.

We also used the Bayesian deterministic approximation of posteriors approach (as implemented in the DAP-G software tool)^53,54^. For each locus, we defined the region of association by identifying the set of variants with LLR<1:1000, ordered the variants based on increasing chromosomal position, and selected the median position to create a +/-500 kilobase (kb) fine-mapping window. Fine-mapping windows were visually inspected manually and adjusted to +/- 1.5 Mb for four loci where 500 kb was insufficient to capture all of the association signal (5p15.3, 16q24.3, 11q14.3, and 20q11.22). The test statistic (Z-score) for each variant from the GWAS summary statistics and the LD matrix (pre-computed using n=∼337,000 unrelated British-ancestry individuals from the UK Biobank^55,56^, s3://broad-alkesgroup-ukbb-ld/UKBB_LD/) were used for the analysis. We set the maximum number of causals at each locus as 5, with exception of the *9p21.3* locus (locus number 27, risk signal numbers 30-35, **Table S1**), where the number of causals was set to six to account for the six independent genome-wide significant signals at this locus. Note, while we allowed for multiple causals/credible sets, for the purpose of fine-mapping we only retained variants within individual credible sets that directly correspond to each of the 68 independent genome-wide significant GWAS signals.

### GWAS conditional analysis

To identify independent risk-associated signals at the *MDM4* locus, we performed conditional and joint association analyses of melanoma GWAS summary data^10^ using the Genome-wide Complex Trait Analysis (GCTA, v1.94.1)^57^ COnditional and JOint association (COJO) module^58^, default settings, and a genomic window of chr1: 203021577-206021577 (hg19). We used an LD reference population of 5,000 individuals selected randomly from the UKBB population determined to be European by PCA (LD_EUR_); variants were converted to best-guess genotype (threshold 0.3) followed by data cleaning for missingness > 3%, HWE p < 1 × 10^−6^, and MAF < 0.001. Separate analyses were performed conditioning on lead variants from credible sets identified via Bayesian fine-mapping (DAPG credible set 1 lead variant: rs2369633, DAPG credible set 2 lead variant: rs12119098). For the conditional analyses, we selected the following genomic window chr1: 203021577-206021577 covering both the DAP-G credible set signals.

### Variant effect prediction

We annotated all fine-mapped variants using Variant Effect Predictor tool (https://grch37.ensembl.org/Homo_sapiens/Tools/VEP)^59^ based on human genome GENCODE version 19 protein coding transcripts. The rsID and “Consequence” columns were extracted and deduplicated to obtain a list of rsIDs and their possible impacts. Additional descriptions of the predicted “consequences” for a given variant can be found at https://useast.ensembl.org/info/genome/variation/prediction/predicted_data.html.

### Melanocyte cell culture

We obtained frozen aliquots of melanocytes, isolated from discarded foreskin tissue of healthy newborn males, from the SPORE in Skin Cancer Specimen Resource Core at Yale University as described previously^23,24^. For capture-C analysis we used three distinct cultures of European ancestry and two of African ancestry (C24, C27, C56, C140, C205). Melanocytes were either grown in Dermal Cell Basal Medium (ATCC PCS-200-030) supplemented with Melanocyte Growth Kit (ATCC PCS-200-041) and 1% amphotericin B/penicillin/streptomycin (120-096-711, Quality Biological) for QTL^23,24^ and capture-HiC analysis, or alternatively in M254 (Invitrogen, M254500) supplemented with HMGS-2 (Invitrogen, S0165) for all other experiments. Cells were grown at 37°C with 5% CO_2_. All cells tested negative for mycoplasma contamination using MycoAlert PLUS mycoplasma detection kit (LT07-710, Lonza).

### Capture-HiC bait design and library preparation

Capture-HiC baits were designed by Arima Genomics (San Diego, CA, 2x tiling, least stringent masking, XTHSBoosting) to obtain an Agilent Sure Select library (Santa Clara, CA) targeting all restriction fragments (recognition sequences: ^GATC, ^GANTC) covering entire regions of association for the 68 independent genome-wide significant signals (**Table S2**)^10^.

For most regions, we used the LLR- and LD-based fine-mapping to define regions of association. The region of association was defined by the two outermost fine-mapped variants and was extended by at least one restriction fragment. Exceptions to this were made for the following genomic loci: For the *5p15.33* locus (locus 11, signals 11-13, **Table S1**), capture baits were designed to cover the entire region spanning both the *TERT* and *CLPTM1L* genes (chr5: 1230000-1360000, ∼130 kb). For the *7q31.11* locus (locus 22, signal 25), we extended the region of association to encompass the complete LD block as defined by LD link ^50-52^ (chr7:124392512-124710858). For the *9p21.3* locus (locus 27, signals 30-35, **Table S1**), we included the entire region spanning from the *MTAP* to *DMRTA1* genes (chr9:21790755-22452478, ∼660 kb). Finally, for the *21q22.3* locus (locus 51, signal 65, **Table S1**), we extended the region to include the previously functionally fine-mapped variant rs398206^60^.

Bait sequences are listed in **Table S2**. Hi-C libraries were generated using the Arima HiC kit (Arima Genomics) and the KAPA HyperPrep kit (KAPA Biosystems) following the manufacturer’s protocol. Briefly, 2-4 million cells were crosslinked, enzyme digested, and ligated. The ligated DNA was reverse-crosslinked, fragmented by sonication, and size-selected for adaptor ligation and library amplification. The HiC library was then hybridized with the custom capture baits and captured by the SureSelect XT HS and XT low input library preparation kit for ILM (Agilent). 15 capture-HiC libraries were made from five human primary melanocyte cultures (C56, C140, C205, C24, and C27) with three technical replicates per melanocyte culture. Barcoded capture-HiC libraries were pooled and sequenced using an Illumina Novaseq, with one run on an SP and a second run on an S1 flowcell, generating ∼5.7 billion paired-end reads with 150bp read length, for a median coverage of ∼350 million read pairs per technical replicate, ∼1.1 billion read pairs per culture.

### Capture-HiC chromatin interaction analysis

Paired-end sequencing reads were pre-processed using the HiCUP pipeline^61^ and aligned to the human genome version 19 using Bowtie2^62^. The summary of quality-control (QC) of sequencing reads for each replicate are shown in **Table S3**. For each melanocyte culture, the aligned reads were pooled across the technical replicates. Chromatin interaction loops were detected at one and four restriction fragment resolutions, separately, using CHiCAGO pipeline version 1.16.0^63^, treating each of the five melanocyte cultures as biological replicates. As described previously^42^, the four-fragment resolution was created using the artificial “.baitmap” and “.rmap” files, where four consecutive restriction digestion fragments were grouped into one fragment (baitmap files provided in **Tables S4 and S5**). We used the default parameters for one-fragment analysis except for minFragLen, maxFragLen, binsize, maxLBrownEst which were set to 75, 1200, 2000, and 150000 respectively (**Table S6**). Four-fragment analysis was conducted using default parameters except for minFragLen, maxFragLen, binsize, maxLBrownEst which were set to 150, 5000, 8000, and 600000 respectively (**Table S6**). Based on the literature^39,42^, for four fragment analysis the removeAdjacent parameter was set to FALSE. Following CHiCAGO tool recommendations, chromatin interaction capture-HiC loops with CHiCAGO scores ≥ 5 were considered high-confidence interactions and were further analyzed. The output file was generated using the long range interaction format and big interact format for visualization on the WashU Epigenome Browser^64,65^ and UCSC genome browser^66-70^ respectively.

Considering the wide possible range of cross-linking resolution, the potential for incomplete restriction digestion, as well as the fact that some variants may be located at the edge of restriction fragment bins, data from adjacent bins may also reflect physical interactions from fine-mapped variants to genes. Therefore, to inclusively identify such interactions, we also considered interaction data from adjacent restriction fragment bins. Specifically, for each fine-mapped variant, we defined a genomic window +/- 500 bases and assessed whether any adjacent restriction fragments overlap this window. In this case, we assessed chromatin interactions from the restriction fragment bin harboring the variant itself as well as any overlapping adjacent restriction fragment bin.

### Capture-HiC target gene nomination for GWAS risk loci

For each GWAS signal, we mapped chromatin interaction loops between baited restriction fragments overlapping fine-mapped risk variants (see above) and the promoter regions of the target gene transcripts as per the GENCODE version 19^67,71^. We defined the promoter regions of the respective target genes using three criteria, identifying genomic regions with histone marks consistent with active promoters in melanocytes and melanoma cells, as well as using a broader definition for gene promoters regardless of activity in melanocytic cells. Specifically:

1. **Melanocyte-specific active promoter regions**: We annotated genomic regions as melanocyte-specific active promoters using the NIH Roadmap Epigenomics Mapping Consortium^16^ ChromHMM imputed state model annotations^72,73^ derived from human primary neonatal melanocyte cultures (imputed ChromHMM state model data was downloaded from the UCSC genome browser Roadmap Epigenomics Integrative Analysis Hub for melanocyte samples E059, E061). Genomic regions were annotated as melanocyte-specific promoter if they overlapped with the ChromHMM imputed model states annotated as: PromU (Promoter Upstream transcriptional start site; TSS), PromD1 (Promoter Downstream TSS with DNase), PromD2, TssA (Active TSS), PromP (Poised Promoter), PromBiv (Bivalent Promoter), Tx_Reg (transcription regulator). Data were available from two melanocyte cultures (E059, E061) and were merged such that regions were defined as promoter if they overlapped promoter annotated sequence from either of the two cultures. Subsequently, target genes were assigned for these promoters based on whether the promoter region overlapped +/-2.5 kb of a transcription start site (TSS) for any GENCODE version 19 comprehensive protein coding and non-coding transcripts^71^.
2. **Melanoma-specific active promoter regions**: Similarly, we defined melanoma-specific active gene promoters using publicly available ChromHMM data from two different engineered melanoma cell-models, HMEL and PMEL^74^. Both HMEL and PMEL cell lines were originally derived from primary foreskin melanocytes, immortalized by overexpression of *TERT*, and introduction of oncogenic *CDK4*^R24C^, dominant negative *TP53*, and *BRAF*^V600E^ ^75^. We used the ChromHMM data specifically from tumorigenic cell line variants with shRNA mediated *PTEN* knockdown (HMEL-sh*PTEN* and PMEL-sh*PTEN*)^74^. Melanoma-specific active promoter regions were defined if annotated as the following ChromHMM states: 1_TssA, 2_PromWkD, 3_TssWkP. Target genes were assigned where the promoter region overlapped +/-2.5 kb of a TSS for any GENCODE version 19 comprehensive protein coding and non-coding transcripts^71^.
3. **Globally defined gene-promoter regions**: We utilized the ENCODE based^76^ promoter boundary criterion in order to more globally define promoter regions regardless of activity in melanocytic cells. Global promoters were defined as regions +/-2.5 kb of a TSS for each of the GENCODE version 19 comprehensive protein-coding transcripts ^71^. 16,663 genes annotated with a respective promoter using the globally defined promoter criteria also have melanocyte- and/or melanoma-specific active promoters.

### ATAC-seq library generation and data analysis

As described previously^47^, 30K-50K primary melanocytes were lysed with cold lysis buffer (10 mM Tris-HCl, pH 7.4, 10 mM NaCl, 3 mM MgCl2, 0.1% IGEPAL CA-630) and centrifuged to obtain nuclei. The nuclei were resuspended in the transposition reaction mix (2x TD Buffer, Illumina Cat #FC-121–1030, Nextera), 2.5 µl Tn5 Transposase (Illumina Cat #FC-121–1030, Nextera) and Nuclease Free H_2_O on ice and then incubated for 30 min at 37C. The transposed DNA was then purified using the MinElute Kit (Qiagen). PCR amplification was performed using Nextera primers for 12 cycles to generate each single library and PCR reaction cleanup was performed using AMPureXP beads (Agencourt). ATAC libraries were sequenced on an Illumina NovaSeq platform using paired-end sequencing. We sequenced 15 ATAC libraries from five independent primary melanocyte cultures (C24, C27, C56, C140, C205), with three technical replicates for each melanocyte culture. The ATAC-seq reads from the technical replicates were merged for each melanocyte culture. We processed the ATAC sequencing (ATAC-seq) data using the ENCODE ATAC-seq pipeline version 1.6.1 (https://www.encodeproject.org/atac-seq/), treating five melanocyte cultures as biological replicates. Sequencing reads were aligned to hg19 using bowtie2^62,77^. The pipeline requires generating two pseudo replicates via random sampling of reads from pooled biological replicates for peak calling. ATAC peaks were called using MACSv2 peak caller (2.1.0)^78^ (*P* < 0.01) and regions overlapping ENCODE blacklisted regions were removed^79^. ATAC peaks were called from the five melanocyte biological replicates, pooled biological replicates, and the pseudo-replicates. The ATAC overlap reproducibility peaks were identified via the optimal criteria assessing peak overlap between individual biological replicates, pooled biological replicates, and the pseudo biological replicates. ATAC-peaks were analyzed and visualized on the WashU Epigenome Browser^64,65^ and UCSC genome browser^66-70^.

### ATAC-seq data from melanoma cell lines

We analyzed publicly available omni-ATAC-seq data^80^ generated from nine melanoma cell lines derived from melanoma patient biopsies^80^. Melanoma cell lines were annotated as belonging to the following melanoma tumor states: melanocyte-like melanoma cell line (data available from three individual cell lines), the intermediate-like melanoma cell lines (data available from three individual cell lines), and mesenchymal-like melanoma cell lines (data available from three individual cell lines). We assessed fine-mapped variant overlap with ATAC-seq peaks in any of the nine melanoma cell lines using the Bedtoolsr package (intersect function)^81,82^.

### Melanocyte-specific and melanoma-specific enhancers

To refine variant and candidate gene selection we utilized publicly available chromatin state data from both melanocytes and melanoma cells to identify genomic regions with promoter or enhancer histone marks. Promoter regions were defined as described above. We annotated genomic regions as melanocyte-specific enhancers using Roadmap ChromHMM data from two primary human melanocyte cultures^16,73^. We utilized the following enhancer annotated states in the primary ChromHMM (Enh, EnhG, EnhBiv), auxiliary ChromHMM (EnhG1, EnhG2, EnhA1, EnhA2, EnhWk, EnhBiv), or the imputed ChromHMM (TxEnh5, TxEnh3, TxEnhW, EnhA1, EnhA2, EnhAF, EnhW1, EnhW2, EnhAc) model data to define melanocyte specific enhancers. We classified a region as enhancer if marked as an enhancer in either cell line for either model. Melanoma-specific active enhancers were defined as regions annotated as any of the following ChromHMM states: 4_EnhA, 5_EnhM, 6_EnhW, 7_TxEnhM, 7_TxEnhW, and 9_TxWkEnhW, from the tumorigenic melanoma cell models described above^74^; regions were classified as an enhancer if marked as an enhancer in any cell line. Any region annotated as enhancer or promoter was considered regulatory. Data were analyzed using the Bedtoolsr package^81,82^.

### Massively Parallel Reporter Assay (MPRA) data from melanocytes and melanoma cell lines

We previously performed episomal Massively Parallel Reporter Assays (MPRA) in immortalized primary melanocytes and the UACC903 melanoma cell line to assess variant allele-specific transcriptional activity^25^. The MPRA study design conducted fine-mapping using LLR- and LD-based criteria similar to the fine-mapping strategies utilized in this manuscript (LLR<1:1000 to the primary lead variant at each locus, LD r^2^>0.8 1000 Genomes Project Phase 3 EUR for secondary signals and variants not included in the GWAS summary data). 1,701 out of the 1,948 fine-mapped candidate variants from this study were successfully tested using MPRA. The remaining 247 variants could not be assessed for two reasons^25^: 102 were not amenable to design or failed assay QC, and 145 were not fine-mapped in the MPRA study due to slightly different fine-mapping criteria.

### QTL colocalization and TWAS/MWAS in melanocytes and melanoma

QTL Colocalization as well as TWAS/MWAS analyses of the GWAS summary data with the expression-QTL or methylation-QTL datasets were available from our previous melanocyte QTL studies^10,23,24^ (dbGaP: phs001500.v1.p1); melanocyte-eQTLs and meQTLs were generated using 106 primary melanocyte cultures derived from individuals mainly of European descent; methylation probes were assigned to genes as previously described (CpG probes located within 1.5 kb of the TSS, 5ʹ-UTR, 1^st^ exon, gene body, or 3ʹ-UTR of a gene)^23^. Colocalization for the secondary marginal GWAS signal near locus 4 (signal 4) was performed using the ezQTL website (https://analysistools.cancer.gov/ezqtl/#/home)^83^ (Parameters: LD-1000 Genomes Project EUR population, window= +/-250 kb). ezQTL performs colocalization using HyPrColoc^84^ as well as eCAVIAR^85^ (eCAVIAR results use a 100 kb window centered on the lead GWAS variant). Consistent with prior studies of melanocyte QTLs^23,24^, we considered colocalization significant where the HyPrColoc posterior probability exceeded 0.5 or the eCAVIAR CLPP exceeded 0.01.

In addition, we assessed QTLs from TCGA melanoma tumors^86^. eQTL colocalization was performed using pre-analyzed QTL data on the ezQTL website. We performed melanoma TWAS using the pre-computed weights from 103 TCGA melanoma samples (http://gusevlab.org/projects/fusion/#the-cancer-genome-atlas-tcga-tumornormal-expression) using FUSION (http://gusevlab.org/projects/fusion/)^87^. Melanoma meQTL colocalization and MWAS were performed as described previously^23^.

For assessing nominal eQTL support for 195 high-confidence candidate genes nominated via chromatin interaction data, we assessed QTLs specifically between the interacting fine-mapped variant and its putative target(s); where candidate genes were outside the +/- 1 Mb *cis*-window previously used for melanocyte eQTL analysis^24^, we specifically tested the association between fine-mapped variant genotype and nominated target gene expression in the melanocyte eQTL data using a linear regression model, where the input to the model included the interacting variant genotype and additional covariates (3 genotyping principal components and 15 PEER factors) from the previous melanocyte study^24^. TCGA melanoma eQTLs were only assessed for genes within +/- 1Mb *cis* window.

### Candidate gene expression in melanocytes and melanomas

For use with an integrative scoring system (described below), we assessed gene expression in melanocyte^24^ and melanoma (The Cancer Genome Atlas project, SKCM Pan-Cancer build accessed via cBioPortal)^86^ gene expression datasets. For each gene expression dataset, we filtered out genes that are not expressed by excluding those with an RSEM value <0.1 in more than 20% of samples criteria. For the remaining genes, we calculated the median expression percentile across samples in the melanocyte and melanoma datasets, respectively.

### Pathway enrichment analyses

Pathway and upstream-regulator enrichment analyses were performed using the Ingenuity Pathway Analysis (IPA) tool (Qiagen)^88^. Pathway enrichment *P*-values were calculated using the Ingenuity knowledge base (genes only) as the reference set and using default parameters. The IPA tool parameter “Relationships to consider” was set to “Direct relationships” for the upstream regulator analysis.

### CRISPRi validation of regulation of target genes transcription

CRISPR interference (CRISPRi) was performed in the immortalized human melanocyte cell line C283T/dCas9-KRAB. The immortalized human melanocyte cell line C283T^47^ was infected with a lentiviral vector pLX_311-KRAB-dCas9 (gift from John Doench, William Hahn, and David Root; Addgene plasmid # 96918; http://n2t.net/addgene:96918; RRID:Addgene_96918)^89^ followed by monoclonal cell selection. We validated dCas9-KRAB expression and activity in the clone used for CRISPRi validation. For each variant tested, three different guide RNAs (gRNAs) were designed to target the genomic regions surrounding the variant, with the gRNA sequence located within/around +/- 50bps from the variant (sequences for guides designed to target the region surrounding each variant are listed in **Table S7**). Two non-targeting gRNAs were used (NTC1, NTC2). gRNAs were ligated into the lentiviral vector pXPR-050 (gift from John Doench and David Root, Addgene plasmid #96925; RRID: Addgene_96925)^90^. Cells were infected with lentiviral particles encoding gRNA and at 24h after infection, 1.5 μg/mL of puromycin was added for selection. After two days of puromycin selection, puromycin was removed, and cells were harvested for RNA collection on the same day or one day after puromycin removal. Total RNA was isolated with RNeasy Mini Kit (Qiagen) and cDNA was generated with SuperScript IV VILO Master Mix (Thermo Scientific). For each variant, at least three infections were performed, with two biological replicates per infection for each of the three gRNAs against the variant. mRNA levels of the candidate target genes being assessed were measured by Taqman assay (Thermo Scientific) and normalized to *GAPDH* levels. qPCR triplicates (technical replicates) were averaged and subsequently considered as a single data point. Data from NTC1 was used for statistical comparisons of other gRNAs, data in tables and graphs were represented as fold-change relative to the average of NTC1. The statistical analysis was performed using a paired two-tailed t-test comparing delta-Ct values.

### Mutational cancer driver genes

Mutational driver genes identified from analysis of melanoma cohorts and pan-cancer dataset were available from the intOGen database^91^ (https://www.intogen.org/search?cancer=MEL and https://www.intogen.org/search#driver-genes:table). We used this nominated target gene list to identify if any of the candidate genes were also identified as cancer drivers in melanoma and pan-cancer datasets.

### Candidate gene prioritization via integrative scoring

We created a scoring scheme (**Figure S1**) for candidate gene prioritization at each melanoma risk signal using complementary information from fine-mapping, chromatin interaction, cell-type specific epigenomic (chromatin state, accessibility), gene expression (eQTL/TWAS), DNA methylation (meQTL/MWAS), MPRA, and mutational cancer driver datasets.

At each risk signal, if a candidate gene was nominated via melanocyte- or melanoma-specific expression QTL colocalization analyses or TWAS, we considered it as a strong plausible candidate and added 6 points to the total score for the gene.

For the remaining genes, we cumulatively assigned scores if:

- The gene is nominated by a fine-mapped variant being linked to the gene promoter via chromatin interaction or physical location within a promoter (+1)
- The gene is nominated by a fine-mapped variant within a melanocyte or melanoma enhancer/promoter region and being linked to the gene promoter (+1)
- The gene is nominated by a fine-mapped variant that overlaps a melanocyte or melanoma enhancer/promoter region, displays significant allelic transcriptional activity, and is linked to the gene promoter (+1)
- The gene is nominated by a fine-mapped variant that overlaps a melanocyte or melanoma enhancer/promoter region, displays significant allelic transcriptional activity, is a marginally significant eQTL for the gene (*P* < 0.05), and is linked to the gene promoter (+1)
- The gene is nominated via methylation colocalization analysis or MWAS approaches (+2).

For all nominated candidate genes, we then added to the score if a candidate gene was identified as driver in melanoma (+1) or pan-cancer (+1) analyses from the intOGen database^91^.

## RESULTS

### Fine-mapping 68 independent melanoma risk signals from GWAS

We performed fine-mapping at 68 genome-wide significant melanoma GWAS risk signals^10^ using a combination of complementary approaches in order to comprehensively and inclusively identify potential causal variants (credible causal variants: CCVs). We first fine-mapped using GWAS summary data, selecting variants with log-likelihood ratios (LLR) <1:100 relative to the lead variant at each locus. We also performed Bayesian fine-mapping using DAP-G^53,54^, identifying credible sets that directly correspond to each of the 68 signals. Lastly, to identify potential causal variants that are not present in the summary data due to quality control filters and or imputation reference choice, we used an LD-based fine-mapping strategy (r^2^>0.8, 1000 Genomes Project, Phase 3, Version 5, EUR). We selected all variants fine-mapped by at least one approach as CCVs, for a total of 1,948 variants. 1,477 variants were fine-mapped by DAP-G, while 1,892 were fine-mapped using the LLR/LD approaches (**Table S8**). 1,421 were identified by both approaches, suggesting that distinct fine-mapping approaches largely prioritize the same set of variants as credible causal variants (**Figure 1A, Figure S2**).

As expected, a large proportion of the fine-mapped CCVs were in non-protein coding regions and only a few variants were identified as directly impacting the protein coding sequence (**Tables S9 and S10**). Among the 1,948 fine-mapped variants, variants altering protein coding sequence (i.e., variant annotated as missense variant or frame shift variant) were observed at the 20 of the 68 GWAS risk signals (20 genes; **Tables S9 and S10**); we observed no variants in consensus splice donors or acceptors. Therefore, at these risk signals we considered the affected gene as a potential target gene based on the variant impacting protein coding sequence; amongst these are well-established pigmentation genes (*MC1R*^92-94^, *SLC45A2*^92,95-97^, *TYR*^2,10,92,98,99^), well-characterized protein-coding melanoma risk variants (*MITF*^100,101^), and variants in well-established cancer genes (*OBFC1*^8,102,103^, *ATM*^104,105^). Most melanoma GWAS risk signals lack protein-altering CCVs, and even still, the presence of such variants within a credible set for a locus does not exclude the possibility of *cis*-regulation being the causal mechanism.

### Mapping chromatin interactions at 68 GWAS signals using a custom region-focused capture-HiC assay

We performed a custom capture-HiC assay to resolve chromatin interaction patterns at melanoma GWAS risk signals. We designed custom capture baits tagging all restriction digestion fragments tiled across the entire region of association for each locus (**Table S5**). 88% of fine-mapped variants (1,717 out of the total 1,948 fine-mapped variants) were located within a baited restriction fragment (**Table S8, Figure S3**). 94% of variants were either directly baited or were located adjacent to at least one baited restriction fragment (**Table S8**). As binning groups of adjacent restriction fragments has been shown to increase sensitivity to detect long range interactions^39,40,47,106^, we also assessed baiting coverage for bins of four adjacent restriction fragments (**Table S5**). We observed slightly better coverage of fine-mapped CCVs in four-fragment analysis, as 95% of variants (1858 out of 1948) were located within a four-fragment bin with at least one restriction fragment that was baited (**Table S8, Figure S3**). The capture-HiC assay was performed using five primary cultures of human melanocytes from unrelated individuals drawn from the melanocyte collection we previously used for eQTL and meQTL studies^23,24^. To maximize sensitivity to detect long-range interactions as others have done, as well as to assess regions harboring variants that were not directly baited, we assessed collective groupings of four adjacent restriction fragments (e.g. four-fragment analysis, 4F; **Table S5**) and compared to the analysis of individual fragments (one-fragment analysis, 1F; settings for both analyses summarized in **Table S6**).

Consistent with previous studies^39,42^, 4F analysis identified chromatin interactions spanning longer distances (**Table S11**) in comparison to the 1F analysis. As expected given the larger number of bins analyzed, the 1F analysis overall identified a higher number of unique chromatin interactions with better resolution (>2.3 fold). We also observed similar distributions of CHiCAGO scores between the 1F (median score=8.38) and 4F (median score=7.07) analyses (**Table S12**). Given the better coverage of variants and increased sensitivity to detect long-range interactions, we used 4F data in the subsequent analyses.

### Capture-HiC links fine-mapped risk variants to candidate target genes at most loci

Next, we analyzed the capture-HiC chromatin interaction data to identify physical interactions between fine-mapped CCVs at the 68 melanoma GWAS signals and gene promoter(s). We defined promoters in three different ways. Firstly, in order to identify promoters in melanoma relevant cell-types where target gene is more likely to be causal, we separately defined melanocyte- and melanoma-specific promoters using ChromHMM state model data from two primary melanocyte cultures (ROADMAP epigenome project^16^), as well as two engineered melanoma cell-models^74^, respectively. We also more broadly defined promoters regardless of activity in melanocytic cells using a general promoter definition from the ENCODE consortium ^69,76,107^.

The capture-HiC data identified chromatin interaction loops from 84% of fine-mapped risk variants (n=1,632) to at least one annotated promoter region, nominating 323 genes as candidate causal genes (CCGs) for 61 melanoma GWAS signals (**Figures 2A-B**, **Figures S4, S5, and S6**). A small proportion of fine-mapped variants were located directly within annotated gene promoters (13%, 263 out of 1,948 variants, 56 unique genes; **Figure 2B**). Most of these not surprisingly showed physical interactions within the promoter itself, however there were nine additional CCVs without such an interaction nominating an additional two genes as potential CCGs (**Figure S7**). 122 promoter-overlapping variants (6%) showed chromatin interaction loops to an alternative promoter for the same gene (located at least 10 kb away). 234 promoter overlapping variants (12% out of 1,948 variants) showed chromatin interactions with gene(s) other than the gene nominated by promoter overlap. Therefore, we considered both chromatin interaction or direct promoter overlap criteria for linking variant to genes and collectively identified a total set of 1,641 unique variants linked to 325 genes (**Figures 2A-B**).

**Figure 2.**
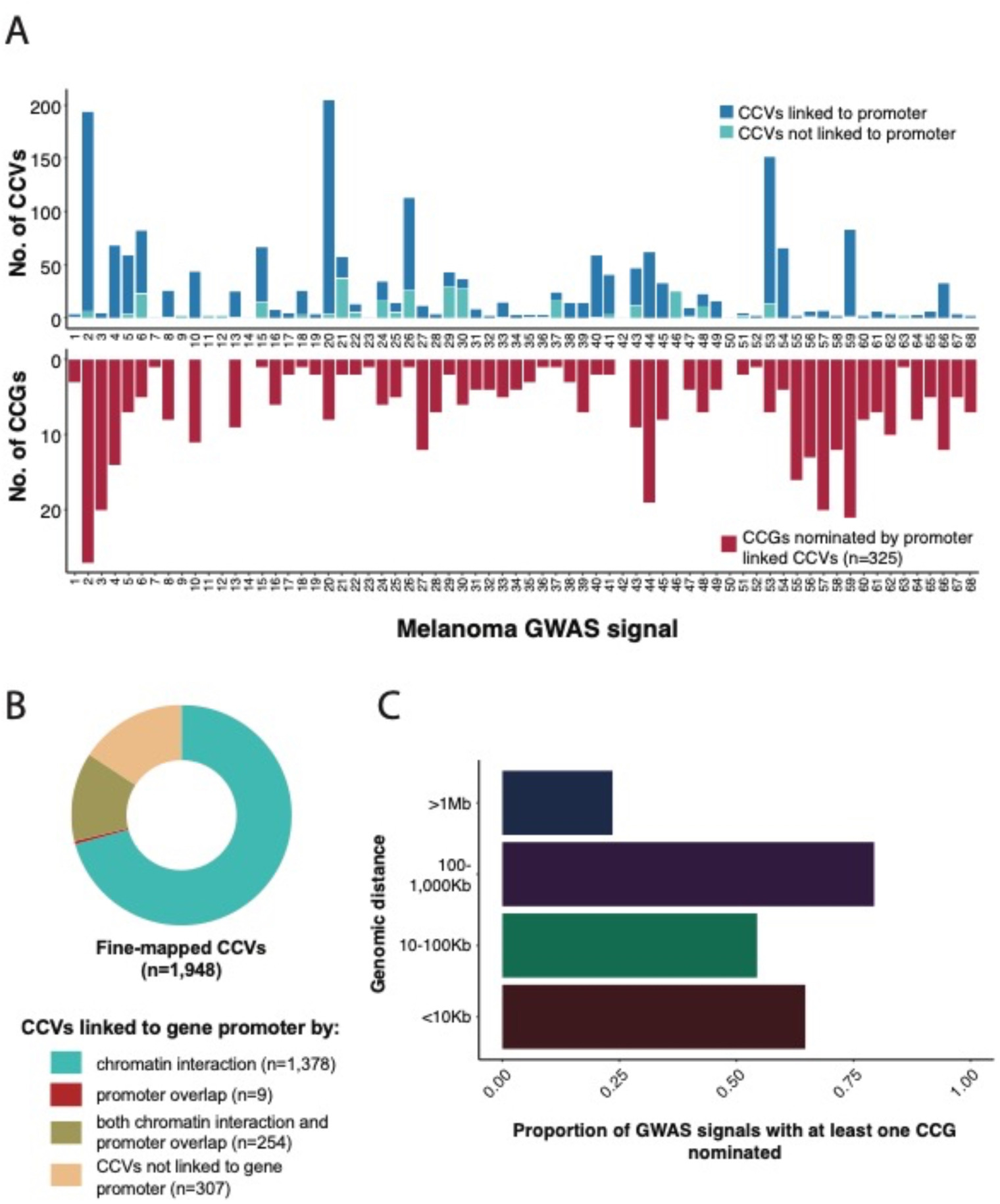
Summary of fine-mapped candidate causal variants (CCVs) linked to potential target candidate causal genes (CCGs) at 68 melanoma GWAS risk signals. (A) Stacked bar plot summary of fine-mapped CCVs and nominated target CCGs. The top bar plot (dark blue color) shows the number of CCVs linked by chromatin interaction or overlap with at least one gene promoter, while the light blue color shows the number of CCVs not linked to a promoter. The bottom plot shows the total number of nominated CCGs per locus. (B) Pie chart showing the proportion of all fine-mapped CCVs that are linked to target CCGs via distant promoter interactions, direct overlap with gene promoter regions, or both. (C) Bar plot summarizing the proportion of GWAS risk signals with at least one gene nominated through chromatin interactions over varying distances.

A median of five candidate target genes were nominated per risk signal; nine GWAS risk signals had only one target gene nominated. Eight fine-mapped CCVs per risk signal (median) were linked to at least one target gene promoter; three risk signals had only one fine-mapped CCV linked to a gene promoter. For seven signals (∼10%), no candidate target could be nominated due to the lack of observed capture-HiC chromatin interaction from the fine-mapped CCVs to any target gene promoter. Of these, three have well characterized pathogenic protein coding changes (signal 9, *MITF* ^100,101^; signal 14, *SLC45A2* ^92,95-97^; and signal 42, *TYR^2,10,92,98,99^*, while three others harbor known melanoma driver or pigmentation genes (signal 11, *TERT*; signal 12, *TERT*; signal 50, *OCA2*;)^5,108-110^. We note that we did not observe the previously reported interaction between rs12913832 within an enhancer in the gene body of *HERC2* and the promoter of *OCA2*^109^.

Notably, we nominated several distant target CCGs via long-range chromatin interaction between fine-mapped CCVs and target gene promoter(s) (**Figure 2C**, **Figure S8**). A large proportion of the GWAS risk signals (79%, or 54/68) had at least one CCG nominated by a chromatin interaction loop spanning between 100-1000 kb distance from the fine-mapped CCV to the target gene promoter (**Figure 2C**). For 23% (16/68) of GWAS risk signals, at least one nominated CCG was located >1 Mb away from the fine-mapped CCV (**Figure S8**).

### Refining variant and gene nomination via integration with cell-type specific epigenomic and massively parallel reporter assay data

Not all interactions between fine-mapped risk variant and gene promoters are necessarily functional *cis*-regulatory interactions, thus we sought to further refine candidate gene nomination using melanocyte- and melanoma-specific epigenomic datasets as well as data from cell-type specific massively parallel reporter assays (MPRA). Firstly, we utilized chromatin accessibility data (ATAC-seq) and chromatin state annotations (ChromHMM) from human melanocytes and melanoma cell lines to identify those interactions between gene promoter(s) and risk variants in potential regulatory elements. We generated ATAC-seq data for the same five melanocyte cultures used in the capture-HiC assay (three replicates per culture) and analyzed treating the five cultures as biological replicates. Additionally, we also utilized publicly available ATAC-seq data from nine melanoma cell cultures^80^. 203 fine-mapped variants (10%) were located within annotated accessible chromatin regions in melanocyte or melanoma cells (**Figure 3A**); of these, 186 variants (9% of 1,948) were linked to a gene promoter via chromatin looping or direct promoter overlap (**Figure S9**, **Table S13**). These variants were linked to the promoter(s) of 223 unique genes nominated as potential candidates at 46 GWAS risk signals. In addition, we performed a similar analysis using melanocyte- and melanoma-derived ChromHMM data, which provides broader regulatory region definitions using multiple histone marks. Specifically, we utilized ChromHMM enhancer and promoter annotations from two human melanocytes cultures from the Roadmap Epigenome Project^16,73^ as well as published data from two engineered melanoma cell models^74^. In contrast to the ATAC-seq data, analysis using ChromHMM data identified considerably more fine-mapped variants located within potentially regulatory regions (n=618, 32%) (**Figure 3A**), with 579 linked via looping or direct promoter overlap to 275 total candidate genes at 56 GWAS risk signals (**Figure S10**, **Table S13**). In total, 610 potentially *cis*-regulatory variant-promoter interactions were identified using either ATAC-seq or ChromHMM data (linked to 282 unique genes, 57 risk signals (**Figure S11**, **Table S13**), while 155 (209 genes) were identified using both datasets. The latter potentially represent stronger functional evidence, however, given functional *cis*-regulatory variants may be found outside of ATAC-seq peaks, we moved forward with a more inclusive approach to identify potential *cis*-regulatory variants (n=610).

**Figure 3.**
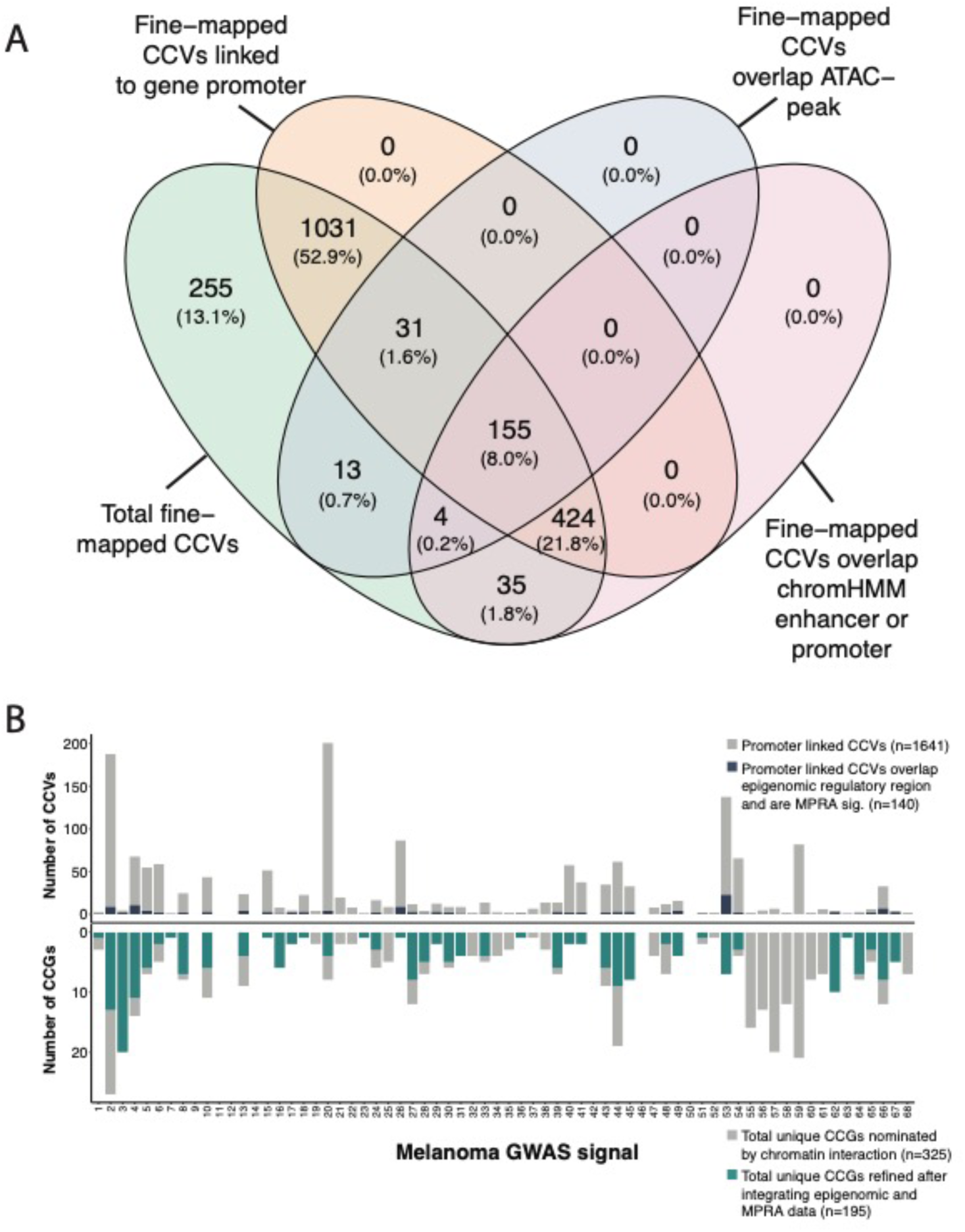
(A) Summary of fine-mapped variants overlap with chromatin interaction *cis*-regulatory regions in the ATAC-seq and ChromHMM datasets. (B) Stacked bar plot summary of fine-mapped variants (CCVs) and nominated target genes (CCGs) after integrating the chromatin interaction dataset with melanocyte- and melanoma-specific ATAC-seq, ChromHMM, and MPRA datasets for each of 68 melanoma risk signals. The top bar plot shows the total number of fine-mapped variants that are linked to at least one target gene using the chromatin interaction dataset, while blue color shows the number of interacting variants overlapping a potential regulatory region in any of the ATAC-seq or ChromHMM datasets and the variant is also FDR significant in MPRA dataset. The bottom plot shows the number of unique genes nominated as potential candidates using chromatin interaction data only, while the green color shows the number of candidate genes following integration with epigenomic (ATAC-seq and ChromHMM) and MPRA datasets.

We then sought to further refine our variant and candidate gene nomination using additional evidence from melanocyte- and melanoma-specific episomal massively parallel reporter assays. We previously assessed allele-specific *cis*-regulatory activity of fine-mapped melanoma risk variants^25^. 1,701 out of the 1,948 fine-mapped variants were assessed in both melanocyte cultures and melanoma cells, of which 349 were FDR-significant in either melanocytes or melanoma cells. We subsequently assessed those MPRA-significant variants located within potentially *cis*-regulatory regions as described above, identifying 140 variants linked to 195 genes at 42 risk signals (“high-confidence gene set”; **Figure 3B**, **Table S13**). While we cannot exclude the possibility that variants and genes filtered out using this strategy may be functional under specific cellular contexts not evaluated here, this set of 140 variants and 195 genes represent strong functional leads.

We looked into cell-type specificity of the high-confidence gene set. In general, roughly half of these high-confidence capture-HiC-nominated genes were identified via interactions with variants that were MPRA-significant and/or located within regulatory regions of both melanocytes and melanomas. Slightly less than half of these high-confidence genes were nominated by analysis of only melanocyte epigenomic/MPRA data but not a similar analysis using melanoma data, e.g. melanocyte-specific candidates (n=88 genes, **Table S14**). Very few gene candidates were identified solely by analysis using only melanoma epigenomic/MPRA data (n=17 genes, **Table S15**). Perhaps unsurprisingly given the role of normal melanocytes in regulating pigmentation, analyses using only melanocyte data nominated candidates for far more loci found to be implicated in pigmentation phenotypes by Landi and colleagues^10^ (n=13 loci) versus analyses using only melanoma data (n=1 locus). Thus, in all, these data suggest that a substantial number of loci may retain function in both melanocytes and melanoma cells and that analyses using melanocyte data appear to better annotate melanoma GWAS loci.

### An integrative scoring system to prioritize GWAS candidate causal genes

We previously used cell-type specific expression and methylation QTL datasets (eQTL, meQTL) to nominate candidate genes for melanoma GWAS risk loci^10,23,24^. Specifically, we performed colocalization for FDR-significant QTLs, as well as identifying FDR-significant transcriptome- and methylome-wide association study (TWAS, MWAS) genes using data from both a panel of primary human melanocyte cultures as well as melanoma tumors from The Cancer Genome Atlas Project^10,23,24^. 40% of candidate genes (22 out of 55) nominated via eQTL colocalization and/or TWAS from melanocytes and melanoma were also nominated as high-confidence genes from capture-HiC data (**Figure 4**, **Table S16**). For meQTL-colocalizing and MWAS-significant genes, only 32% (32 out of 100) were identified in the high-confidence gene set. Of the meQTL/MWAS-nominated candidate genes that lack significant eQTL/TWAS support, 26% (19 out of 74) were nominated as high-confidence candidates in chromatin interaction analyses.

**Figure 4.**
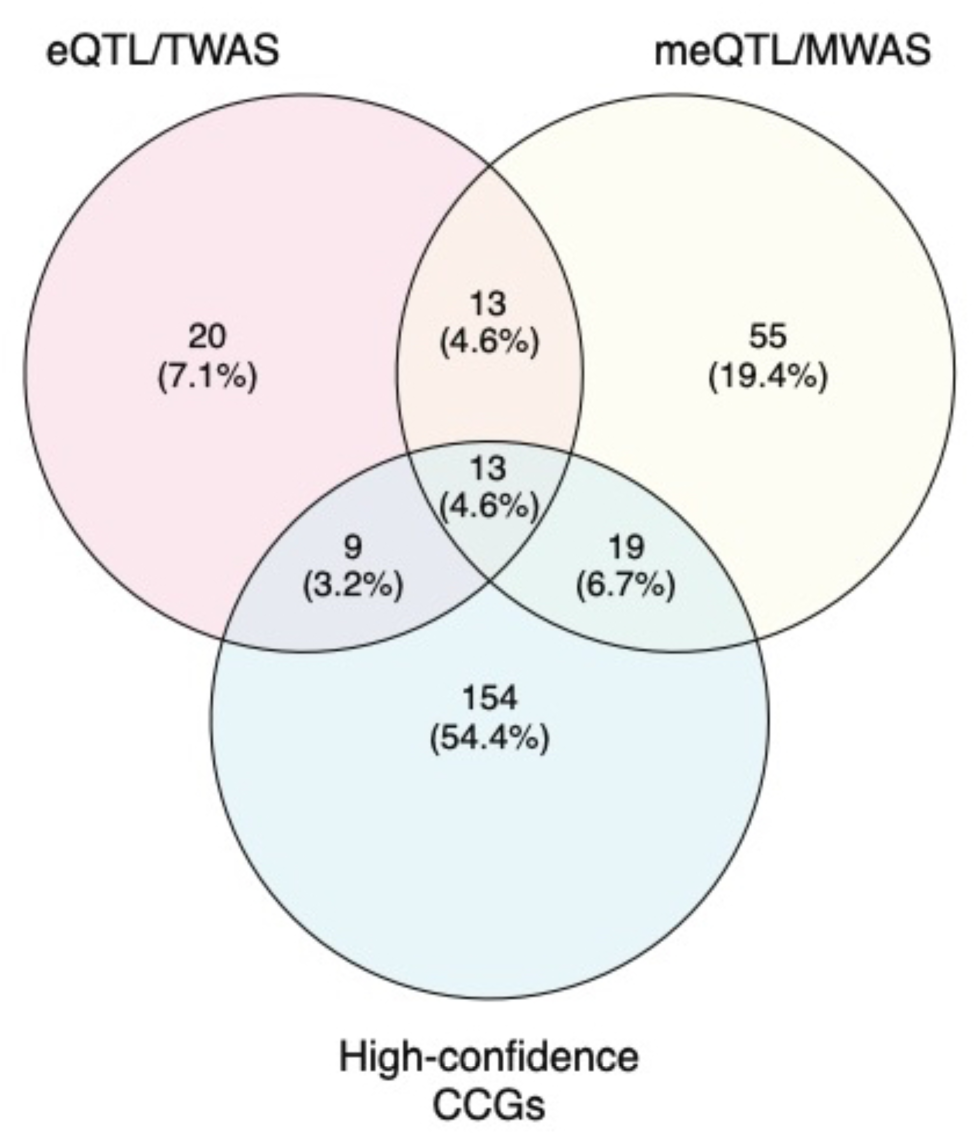
Summary of overlapping CCGs between QTL datasets and capture-HiC chromatin interaction analyses. eQTL/TWAS CCGs were nominated when colocalization of eQTL and GWAS data was observed, or alternatively when the gene was identified as FDR-significant via Transcriptome Wide Association Study (TWAS), using either primary melanocyte or melanoma tumor eQTL reference datasets. Likewise, meQTL/MWAS CCGs were nominated via meQTL colocalization or an FDR-significant Methylome-Wide Association Study finding, where the significant CpG probe was located within a gene promoter or gene body, and meQTL reference datasets from melanocytes and melanoma tumors were tested separately. High confidence CCGs were nominated via integration analyses of fine-mapping, chromatin interactions datasets with epigenomic (ATAC-seq and ChromHMM) and MPRA data derived from melanocytes and melanoma cells.

Given that a large proportion of the 195 genes nominated via chromatin interaction analyses lack significant eQTL/meQTL/TWAS/MWAS support, we assessed whether any of these genes may be supported via marginal eQTLs between the promoter-interacting variant and its putative target(s). In total, 36 such genes displayed at least nominal QTL support (*P* < 0.05) in melanocyte and/or TCGA eQTL datasets (**Table S13**).

Finally, to better prioritize *cis*-regulatory functional leads at each risk signal, we created a candidate gene prioritization score integrating melanocyte and melanoma functional datasets (**Figure S1**). At each signal, we considered the presence of significant colocalizing eQTL or TWAS findings, which suggest potential shared causal variants between gene expression and melanoma risk, to be strong evidence for candidate genes, contributing a total of six points to the gene score. For the remaining gene candidates, we combined fine-mapping, chromatin interaction, cell-type specific epigenomic (chromatin state, accessibility), DNA methylation (meQTL/MWAS), and MPRA evidence to assign a gene score up to six points. For all candidates, we further added one point each where genes have been identified as melanoma or pan-cancer driver genes in the intOGen database^91^, allowing for a maximum gene score of 8 (**Table S17**, **Figures 5A-B**, **Figures S12A-H**). In total, six risk signals had at least one candidate gene score ≥ 7, 37 with a gene scoring ≥ 6, and 49 with a score ≥ 4. At previously characterized loci, the scoring system ranked the likely casual as the highest scoring gene, including *PARP1* at 1q42^111^ (locus 5, score = 6), *AHR* at 7p21^47^ (locus 23, score = 4), and *MX2* at 21q22^60^ (locus 65, score = 6) (**Figure 5A**). For novel loci, the scoring system ranks *MDM4* as the best scoring and *CBL* as the second-best candidate for loci on chromosome bands 1q32 (locus 4, score =7) and 11q23 (locus 44, score =4) respectively. Both genes are located more than 500 kb and 1 Mb from their respective lead variants (**Figure 5B**). In addition, the scoring system nominates two SRY-related HMG-box genes, *SOX4* and *SOX6*, as the best candidates at 6p22 (loci 18-19, scores =4,2) and 11p15 (locus 40, score=6), respectively (**Figure 5B**). Collectively, our integrative scoring system-based gene prioritization approach re-identified previously characterized susceptibility genes and provide additional support for functional investigation of novel candidates.

**Figure 5.**
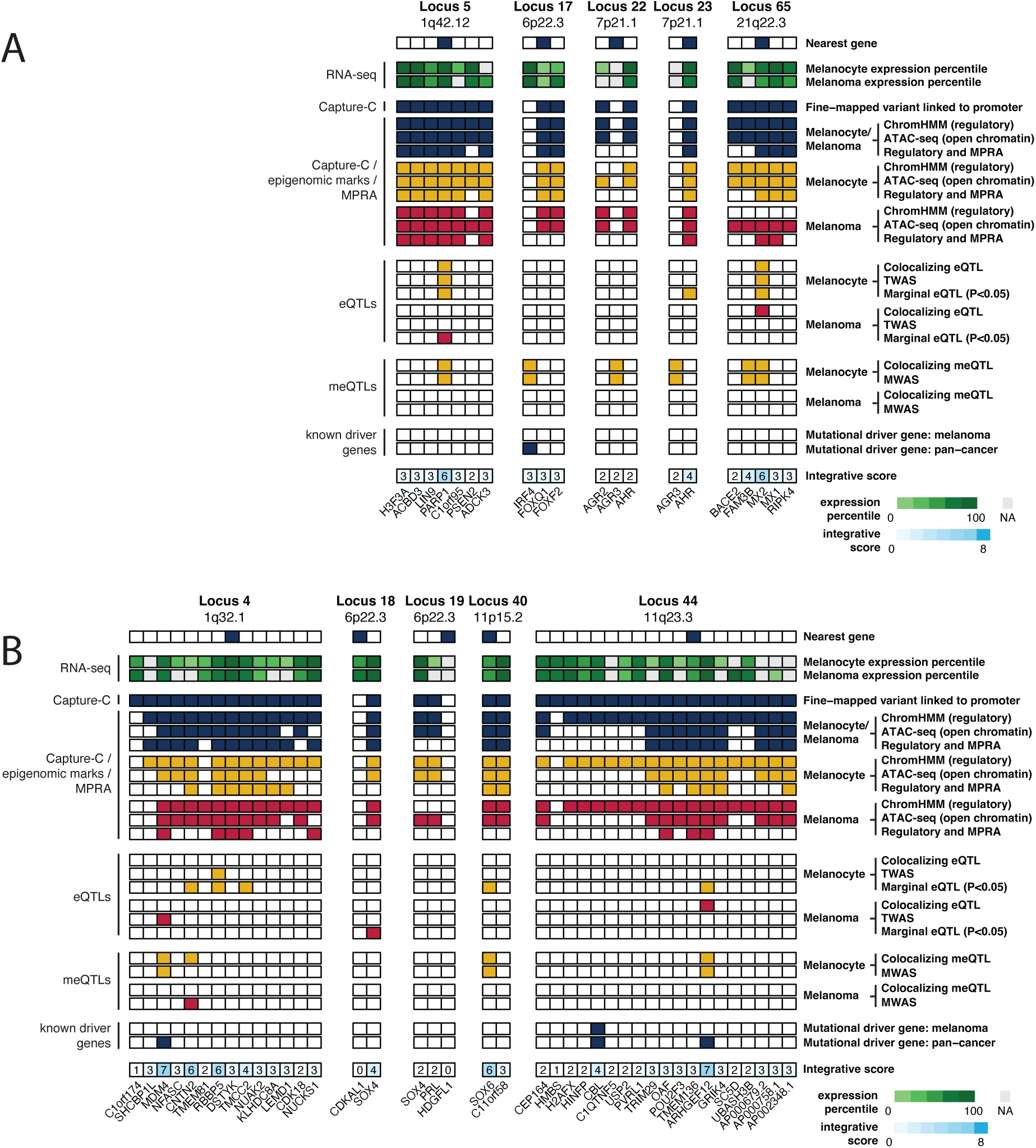
Integrative evidence for candidate causal genes at select melanoma risk signals. (A) Loci with previously characterized candidate causal genes, and (B) select novel loci. For each locus, the figure indicates the nearest gene to the lead variant, summarizes candidate gene expression in primary melanocytes and melanoma tumors, indicates genes implicated by interaction of fine-mapped variants to the gene’s promoter, along with further refined evidence for these interacting variants integrated with melanocyte and melanoma epigenomic and MPRA data. Also summarized are melanocyte eQTL/TWAS evidence, meQTL/MWAS evidence, and whether the candidate gene has been implicated as a melanoma or pan-cancer driver gene. Finally, the figures show an overall integrative score for each candidate scored from 0-8 with 8 being the highest score.

### Biological pathway enrichment analysis of capture-HiC nominated gene candidates

To identify biological pathways associated with melanoma risk, we performed pathway enrichment analysis of the collective set of genes nominated by (1) identification of protein-coding variants amongst the full set of fine-mapped variants, (2) colocalizing eQTLs/meQTLs or FDR-significant TWAS/MWAS findings, and (3) the set of high-confidence gene candidates (n=195 genes) nominated via capture-HiC analyses (**Table S18**). We compared this to a pathway analysis of only these genes nominated by prior QTL (eQTL/meQTL colocalization, TWAS/MWAS) analyses or protein-coding fine-mapped variants (**Table S19**). Overall, pathway analyses including the high-confidence capture-C nominated genes had a greater number of pathways enriched in comparison to that of protein-coding/QTL nominated genes. Novel pathways identified only by the former analysis include p53 signaling (*P* = 0.0005), WNT/beta-catenin signaling (*P* = 0.002), and interferon gamma signaling (*P* = 0.003) (**Table S18**); the capture-HiC gene set further strengthened the evidence for enrichment in numerous pathways, including MITF-M-dependent gene expression (-log10*P* = 4.26 vs 2.87), telomere maintenance (-log10*P* = 3.52 vs 1.9) and aryl hydrocarbon receptor signaling (-log10*P* = 3.22 vs. 1.87; **Tables S18-S19**) along with pigmentation related pathways (melanin biosynthesis, -log10P = 7.28 vs 5.92; L-dopachrome biosynthesis, -log10P = 3.99 vs. 2.0; melanocyte development and pigmentation signaling 3.29 vs. 1.88; **Tables S18-S19**) which is consistent with skin pigmentation-related phenotypes as critical risk factors for melanoma. We also performed enrichment analysis for upstream transcriptional regulators of genes collectively nominated by capture-HiC, eQTL/TWAS, meQTL/MWAS, and protein-coding fine-mapped variants (**Table S20**). We found MITF to be the most enriched upstream regulator, consistent with its well-established role in pigmentation and melanoma risk and progression. In addition, multiple upstream regulators related to MITF were identified as enriched with this gene set, including ZEB2, which has been itself shown to regulate *MITF* levels^112^, as well as SMARCA4 (BRG1) which has previously been shown to be required for MITF activation of melanocyte-specific target genes^113^.

### CRISPRi validation of long-range cis-regulatory interactions

To further assess and validate the regulation of potential target genes by fine-mapped functional variants nominated by integrative analysis, we performed CRISPRi experiments to test the transcriptional regulation of four nominated high-confidence candidate genes at five independent loci. Firstly, we assessed two SOX family transcription factors given a well-established role for SOX proteins in neural crest and melanocyte development, as well as the fact that capture-HiC data identify chromatin interactions with multiple SOX genes. Specifically, *SOX4* was nominated via long-range interactions from two independent loci located more than 1.5 Mb apart (locus 16, signal 18, ∼400 kb; locus 17, signal 19, ∼1.1 Mb, **Figure 6**). *SOX6* was nominated via ∼130 kb chromatin interactions (locus 32, signal 40) and was also identified in melanocyte methylation QTL colocalization and MWAS analyses (**Figure S13A**). In addition, we assessed loci interacting with known cancer driver genes. Specifically, *MDM4* was nominated by long-range chromatin interaction (locus 4, signal 4, ∼550 kb) and was further nominated by melanocyte MWAS and TCGA melanoma TWAS analyses (**Figure S13B**). Finally, *CBL* was nominated by a long-range ∼1.1 Mb interaction (locus 36, signal 44; **Figure S13C**). As eQTLs for *CBL* were not previously been tested^10^ given the 1.1 Mb distance, we evaluated multiple fine-mapped variants and observed a marginal correlation between risk allele and *CBL* expression in primary melanocytes (including rs2120430, *P* = 0.008; rs61898347, *P* = 0.009; rs11217853, and *P* = 0.02; rs61900794, *P* = 0.02; **Figure S14**; **Table S21**).

**Figure 6.**
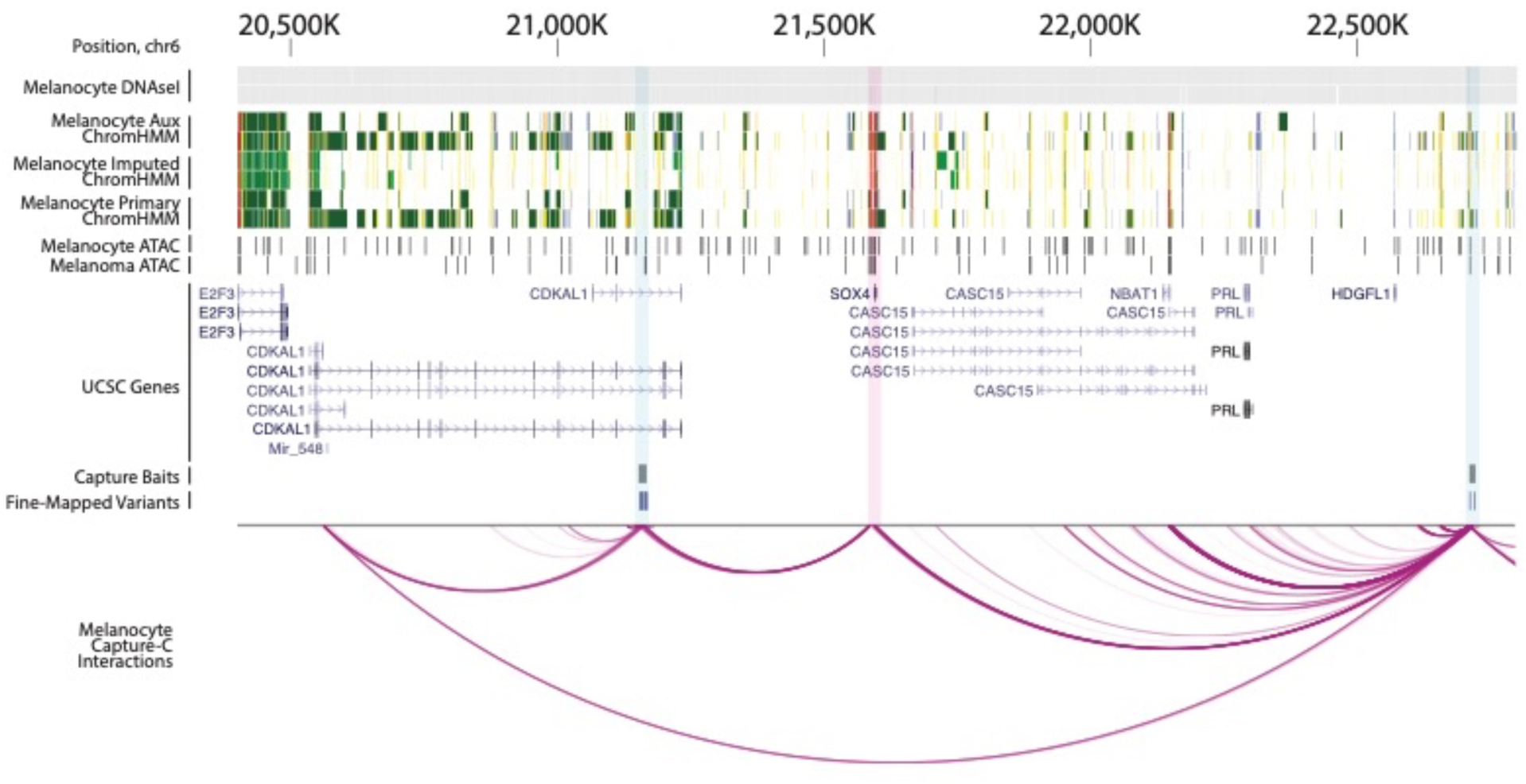
Chromatin looping from two independent loci on chromosome 6 to the promoter of *SOX4*. Figure shows data from melanocyte DNase I hypersensitivity sequencing (Roadmap, n=2 melanocyte cultures), melanocyte ChromHMM (Roadmap, n=2 melanocyte cultures), melanocyte ATAC-seq (n=5 cultures), and melanoma cell ATAC-seq relative to genes in the region. Fine-mapped variants for both loci and location of capture-HiC baits is shown along with chromatin looping. Fine-mapped variants from both loci located within the *CDKAL1* gene and near *HDGFL1*, respectively, directly interact with the *SOX4* promoter region.

For each locus, we chose fine-mapped variants using the collective evidence from capture-HiC, epigenomic, and MPRA datasets, designed three guides targeting the region surrounding each variant, and tested these guides for effect on target gene expression relative to a non-targeting control gRNA (NTC1) in an immortalized human melanocyte cell clone stably expressing dCas9-KRAB.

At locus 16 (signal 18) within the *CDKAL1* gene, we tested four regions harboring a set of five fine-mapped variants within a ∼3 kb stretch (**Figure 7A**); at least one gRNA targeting each of the four CCVs showed significant reduction of *SOX4* without affecting *CDKAL1*. All three gRNAs targeting both rs7776158 or rs2125570 showed significant reductions of *SOX4* (0.63-0.68-fold and 0.67-fold expression relative to non-targeting guide 1, NTC1), as did two out of three gRNAs simultaneously targeting both rs6935117 and rs6935124 (0.72-0.84-fold expression relative to NTC1; **Figure 7B**, **Table S22**). Only one of three gRNAs targeting rs6914598 showed inhibition of *SOX4* (0.72-fold expression relative to NTC1; **Figure 7B**, **Table S22**). None of the guides targeting any of these variants significantly influenced *CDKAL1* (**Figure 7C**, **Table S22**). While further characterization is necessary to disentangle the exact functional variant or combination thereof, these data clearly indicate specific transcriptional regulation of *SOX4* via this risk locus. At a second locus (locus 17, signal 19) closest to the *HDGFL1* gene, we targeted three regions harboring four fine-mapped variants (**Figure 7A**, **Table S22**). While guides targeting rs16886790 showed no significant reduction of *SOX4* levels, all three gRNAs simultaneously targeting both rs72834823 and rs72834822, as well as those targeting rs6456503 significantly reduced *SOX4* levels (0.72-0.80 and 0.75-0.78-fold expression relative to NTC1, respectively; **Figure 7B**, **Table S22**), consistent with the potential regulation of the *SOX4* gene by the region. In contrast to rs1688670, rs72834823, rs72834822, and rs6456503 do not directly overlap with a restriction fragment bin interacting with *SOX4* nor are located within ATAC open or ChromHMM enhancer regions but are in close proximity to interacting enhancer regions. These CRISPRi data nonetheless suggest the regions harboring these SNPs directly regulate *SOX4* expression.

**Figure 7.**
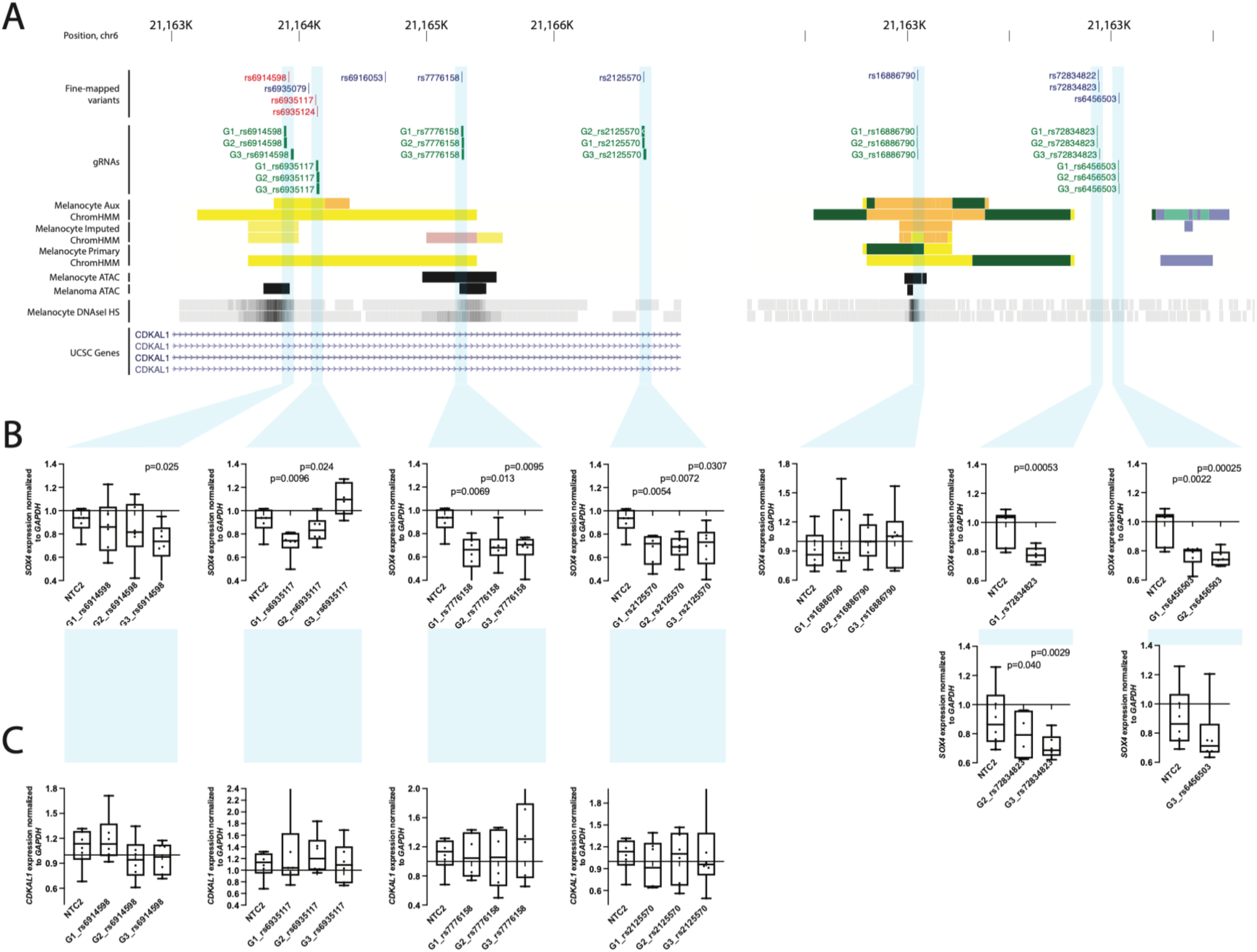
CRISPR-inhibition validation of *SOX4* as a target of regulatory regions harboring fine-mapped variants at two independent melanoma risk loci on chromosome 6. (A) Guide RNAs were designed to target four regions collectively harboring five fine-mapped sequence variants in a risk locus located within an intron of the *CDKAL1* gene (left), as well as three regions harboring four fine-mapped variants for an independent locus nearest the *HDGFL1* gene (right); three guides were designed per region and tested along with two non-targeting guides (NTC1 and NTC2). (B) Each guide was individually tested for effects on *SOX4* expression relative to NTC1 in immortalized melanocytes stably expressing dCas9-KRAB via a TaqMan quantitative RT-PCR assay. Expression values from six replicate experiments are shown as fold change relative to NTC1. Where SNP-targeting guides were tested in separate experiments, they are shown grouped with respective values for NTC2 from the same experiments. Whiskers show minimum and maximum values. *P*-values were calculated using a two-sample two-sided paired t-test comparing delta-Ct values from individual guides to those from NTC1.

For *SOX6* (locus 32, signal 40), we tested five regions harboring six SNPs (rs1455114/rs1455115, rs2953060, rs4617548, rs7108091 and rs7941496). Only one of the five regions (targeting both rs1455114 and rs1455115; interact with *SOX6*; located within melanocyte enhancer; within ATAC-open region in melanocytes and melanoma) showed a reduction of *SOX6* by a single guide (0.71 fold expression relative to NTC1; **Table S22**). A second region targeting rs7108091 (interacts with *SOX6*, MPRA-significant in melanocytes and melanoma) showed marginal reductions for two gRNAs (**Table S22**). On the other hand, at least one guide targeting each of the five regions showed a reduction in *C11orf58*, another interacting candidate gene ∼650 kb from the GWAS signal. All three guides each respectively targeting rs2953060 or rs4617548 showed significant reductions in *C11orf58* (0.34-0.56 and 0.72-0.81 fold-expression relative to NTC1, respectively; **Table S22**), suggesting this gene as a strong candidate causal gene. For *CBL* (locus 36, signal 44), two out of three gRNAs for one of four tested SNPs (rs61900794; melanoma and melanocyte enhancer, ATAC open in melanoma) showed small but significant effects on *CBL* transcription (0.81-0.84 fold expression relative to NTC1). We note that while the restriction fragment bin harboring rs61900794 was not found to interact directly with *CBL* promoter, it is located within a contiguous enhancer region that does, consistent with the region surrounding rs61900794 functioning as an enhancer for *CBL*. Finally, for *MDM4* (locus 4, signal 4), all three gRNAs for one of the four SNPs tested (rs6700182; interacts with *MDM4*; within melanocyte and melanoma ChromHMM regulatory region; MPRA-significant in melanoma) showed significantly reduced *MDM4* levels (0.82-0.85-fold expression relative to NTC1; **Table S22**); only one of these variants reduced expression of another candidate in this region, *RBBP5*, with no effect on a third candidate, *TMEM81*.

### Additional evidence for MDM4 as a melanoma predisposition gene

Bayesian fine-mapping of the locus for which *MDM4* was nominated as a potential causal gene (locus 4, signal 4, lead SNP rs2369633) identified a second set of credible causal variants (**Table S23**; variant with highest posterior inclusion probability is rs12119098). This second credible set appears to be an independent GWAS signal of marginal significance marked by rs12119098 (GWAS *P* = 1.30 x 10^-7^, rs12119098-G OR = 0.95; LD to rs2369633, r^2^ = 0.0016, D’ = 0.18) which remains strongly significant after conditioning on the lead SNP for locus 4 (signal 4, rs2369633; *P*_conditional_ = 2.17 x 10^-7^). In addition, rs12119098 is a significant colocalizing melanocyte eQTL for *MDM4* (*P* = 5.83 x 10^-6^; HyPrColoc posterior probability = 0.78; eCAVIAR CLPP = 0.073; **Figure S15**) but no other gene, where the risk-associated allele is associated with lower *MDM4* levels. rs12119098 is not an eQTL for *MDM4* (*P* = 0.10) or any other gene in melanoma tumors. Taken together with the capture-HiC and CRISPRi already linking locus 4 to regulation of *MDM4*, these reinforce a potential role for *MDM4* in melanoma risk.

## DISCUSSION

With ever growing GWAS sample-sizes for GWAS resulting in increasing numbers of genome-wide significant loci, high-throughput analyses are critically needed to link risk variants with their respective target genes. Most common trait-associated loci identified by GWAS harbor risk variants located primarily in non-coding regions, with the underlying causal variants largely hypothesized to function via altering expression of causal genes. Here, we report a post-GWAS follow-up study aimed at identifying potential causal genes underlying common melanoma risk loci by evaluating cell-type specific chromatin interactions using a custom, GWAS region-focused chromatin capture-HiC assay in human primary melanocytes.

Where eQTL colocalization and TWAS using a primary melanocyte expression reference dataset previously nominated candidate genes for only roughly 25% of the loci from the most recent (2020) melanoma risk GWAS^10^, our GWAS region-focused capture-HiC data identified fine-mapped risk variants either interacting with or overlapping with gene promoters for 61 out 68 risk signals, with a median number of five candidate genes per locus. These data suggest that region-focused capture-HiC assays are highly sensitive for identifying variant-to-gene promoter associations, but alone are not likely sufficiently specific to pinpoint the individual causal gene or genes at many loci. We applied multiple additional filters to narrow down the number of interactions to retain (1) only those involving fine-mapped variants located within cell-type specific (melanocyte or melanoma cell) *cis*-regulatory regions (nominates 282 genes at 57 risk signals), and (2) subsequently requiring these interacting variants to have been significant in a large-scale parallel reporter assay screen conducted in both melanocytes and melanoma cells^25^. This resulted in prioritization of 140 *cis*-regulatory variants interacting with 195 genes at 42 signals, with at least one gene nominated at 62% of risk signals.

Our analysis appeared to be highly sensitive for identifying long-distance interactions. Most risk signals (54/68) had at least one gene nominated by an interaction ranging from 100 kb to 1 Mb. Notably, roughly a quarter of GWAS signals (16/68) had a fine-mapped variant interacting with a gene more than 1Mb away, distances conventionally ignored in *cis*-eQTL analyses. Indeed, as an example we identified multiple fine-mapped SNPs interacting with the promoter of *CBL* that were in fact previously-untested marginal eQTLs for *CBL* (locus 36, signal 44, rs2120430, *P* = 0.007; rs11217853, *P* = 0.02), consistent with potential long-range allelic *cis*-regulation of *CBL* by risk-associated variants. We focused our CRISPRi-based validation efforts primarily on validating longer-range *cis*-regulatory interactions with strong causal candidates, validating such regulatory interactions for two independent loci and *SOX4* (loci 16 and 17, signals 18 and 19, located ∼400 kb and 1.1 Mb away from *SOX4*, respectively), one with *MDM4* (locus 4, signal 4, ∼550 kb distance), and one with *CBL* (locus 36, signal 44; 1.1 Mb distance). Of the loci we tested, we only failed to validate *cis*-regulation of *SOX6* (locus 32, signal 40), but instead showed strong regulation of a distant candidate (*C11orf58*) located more than 500 kb from the risk signal.

*SOX4* was nominated as a potential causal gene via variant-to-promoter looping for two loci originally considered independent based on distance (∼1.6 Mb) and lack of LD, highlighting the limitations of assigning loci in this manner. For the first locus, located within the gene body of *CDKAL1*, we identified three variants each of which physically interact with *SOX4*, show allelic *cis*-regulatory potential via MPRA where the risk allele is associated with higher reporter expression, and are located within CRISPRi-validated *SOX4* regulatory regions (locus 16, signal 18, rs6935117, rs6935124, and rs2125570). In addition, all three of these variants are marginal eQTLs for *SOX4* in TCGA melanomas (rs6935117, *P* = 0.01; rs6935124, *P* = 0.01; and rs2125570, *P* = 0.01; **Table S21**) with direction of effect matching reporter assay data, i.e. risk alleles for these variants are associated with higher *SOX4*. We likewise see physical interaction between fine-mapped variants for the locus near *HDGFL1* and *SOX4* (locus 17, signal 19); while none showed clear allelic regulatory potential via MPRA or were marginal QTLs (**Table S21**), CRISPRi validated regulation of *SOX4* from regions harboring these fine-mapped variants. Collectively, these data provide strong evidence establishing *SOX4* as a potential melanoma risk gene. *SOX4* plays a pivotal role in regulating stemness, promoting cell survival, and epithelial to mesenchymal transition^114-118^. Single-cell sequencing studies have identified *SOX4* as a marker of multiple melanoma cell states including a melanocytic state in human tumors^80^, and an invasive-like melanoma program in patient derived xenografts under RAF/MEK inhibition^119^. Intriguingly, analysis of a zebrafish model found *SOX4* to be a marker of stress-like cell population that more efficiently seeded new tumors; induction of a stress-like program conferred resistance to both BRAF and MEK inhibition in zebrafish melanoma cells^120^. We do not observe a correlation between risk variant genotype and expression of *SOX4* in melanocytes. We did observe at least a marginal correlation with*SOX4* expression in melanomas for one of the two signals, suggesting that the function of causal variants in these two regions may be context-dependent, e.g. dependent on cell state, differentiation, or requiring oncogenic mutations. Beyond *SOX4*, we provide weaker evidence for a second SRY-box transcription factor, *SOX6*. Specifically, while we observe interaction between multiple fine-mapped variants and *SOX6* promoter, CRISPRi validation for the few variant regions we tested did not firmly establish a regulatory link between risk variants and *SOX6*. *SOX6* was recently found to be a marker of a hypothesized intermediate melanoma cell state^80^ between melanocytic cells and a mesenchymal-like state associated with increased migration and resistance to therapies.

These data also provide strong evidence linking two risk loci to distant established melanoma driver genes. We find physical associations between multiple fine-mapped variants (locus 4, signal 4) and *MDM4* amongst other candidate genes. On one hand, rs6700182 is located within a CRISPRi-validated *MDM4* regulatory region and shows allelic regulatory potential via MPRA with the risk allele associated with higher reporter expression (**Table S21**). In contrast a second variant with two CRISPRi guides showing significant or marginal knockdown of *MDM4* (**Table S22**), showed significant allelic regulation in the opposite direction via MPRA (**Table S21**). Fine-mapping of this locus identified a second independent-but-marginal melanoma GWAS signal over the *MDM4* gene itself (rs12119098, *P* = 1.30 x 10^-7^; **Figure S15**), a signal that colocalizes with a significant melanocyte *MDM4* eQTL (*P* = 5.83 x 10^-6^; **Figure S15**), considerably strengthening the evidence for a role for *MDM4* in melanoma susceptibility and potentially resolving a role for *MDM4* in promoting or alternatively protecting against melanoma. Here, we observe a positive correlation between the rs12119098-protective allele and *MDM4* expression in melanocytes (as well as many other tissue types assayed by The Genotype-Tissue Expression (GTEx) project^121^), suggesting higher *MDM4* expression is protective. In contrast to melanoma risk, *MDM4* is found to be amplified in ∼5% of melanoma tumors ^86,122^, is over-expressed in much larger proportion (65%) of melanomas^123^, and antagonizes TP53 function^123,124^. These data suggest potentially pleiotropic roles for *MDM4* across different stages of melanomagenesis; on one hand higher expression of *MDM4* protects against melanoma development while on the other overexpression is selected for during tumor progression and promotes melanoma cell survival. We note that a previously-published small pooled CRISPR knockout screen in melanocytes^47^ found that *MDM4* knockout significantly reduced melanocyte viability and/or growth (FDR = 0.000102), and both RNAi and CRISPR screen data from the Cancer Dependency Map project^125^ show *MDM4* knockout to be strongly selective in the same direction. This locus appears to be potentially pleiotropic in terms of cancer risk; rs12133735 near *MDM4* has been reported as a suggestive association for aerodigestive squamous cell cancers (rs12133735 LD to rs12119098 r^2^ = 0.82), where the rs12133735-G risk allele is on a shared haplotype with the melanoma rs12119098-G protective allele^126^. Further work will be required to understand this pleiotropy and the mechanistic role of *MDM4* in risk. More broadly, pathway analysis of high-confidence candidate genes suggest enrichment for genes involved in p53 signaling including *MDM4* including *TP53* itself (loci 60 and 61, integrative scores of 3 and 4, respectively).

For locus 36 (signal 44), we observe multiple interactions between fine-mapped risk variants and the *CBL* promoter and verified *cis*-regulation of *CBL* by a region harboring at one such variant (rs61900794) via CRISPRi. While this variant did not show significant allelic *cis*-regulatory potential via MPRA, fine-mapped variants at this locus are indeed eQTLs for *CBL* in melanocytes (*P* = 0.02 to 0.008 for the four variants targeted by CRISPRi; **Figure S14**; **Table S21**), suggesting the potential for shared causal variants between melanoma risk and germline regulation of *CBL* expression at this locus. *CBL* plays a role in downregulation of receptor tyrosine kinase signaling including through the RAS-MAPK pathway. Germline *CBL* mutations, primarily missense mutations confined to the linker and RING domains, have been found to be associated with cancer including juvenile myelomonocytic leukemia^127^ as well as a variable syndrome overlapping with Noonan syndrome^128,129^. Somatically, *CBL* has been identified as a potential melanoma driver^91^, with loss of function alterations specifically enriched in desmoplastic melanomas (11%)^130^. While our analysis does not identify one or more clear-cut causal sequence variant candidates, the physical and gene-regulatory connection from this risk locus as well as melanocyte eQTL for *CBL* nonetheless establishes regulation of *CBL* from this locus and suggests that a potential role for *CBL* and common variation underlying RAS-MAPK signaling in melanoma risk should be further explored.

Recently Pudjihartono and colleagues performed an integrative analyses of the 2020 melanoma GWAS risk signals to nominate likely causal variants-target gene pairs using keratinocyte and melanoma genome-scale HiC data, melanocyte and melanoma-specific epigenomic (promoter, enhancer histone marks, and DNA accessibility), and melanocyte and GTEx skin tissue gene expression datasets^131^. While these studies share similarity in approach, there are several key differences. Pudjihartono and colleagues relied on genome-scale Hi-C data generated in keratinocytes and melanoma cells, while we applied a higher-resolution capture-HiC approach to primary melanocytes coupled with deep library sequencing, potentially allowing for more sensitive assessment of interaction in the cell-type of origin of melanoma. We further sought to take advantage of the resolution of this approach by requiring interactions between fine-mapped variants and annotated gene promoters, rather than including interactions with the gene body. Finally, we focused on integrating data specifically from melanocytic cells (melanocytes or melanomas/melanoma cells) and were able to further refine our candidate gene nomination by taking advantage of a larger massively parallel reporter assay which assessed the vast majority of fine-mapped variants from this study for allelic cis-regulatory potential in melanocytes and melanomas. Comparing the high-confidence set of candidate genes identified here, 44 of the 151 genes identified by Pudjihartono were likewise found amongst our 195 gene high-confidence gene set (**Table S23**). These approaches are complementary; inclusion of keratinocyte data and skin eQTLs has the advantage of potentially identifying causal variants and genes that function via gene regulation in a cell type other than melanocytes, while our approach is highly focused on identifying genes that function in a cell-type intrinsic manner. We anticipate both approaches to be of considerable utility as applied to future melanoma GWAS.

We also acknowledge several limitations of our current study. We performed the capture-HiC assay in primary melanocytes, the cell-of-origin for melanoma, and our downstream analyses integrating epigenomic and MPRA data focused on integrating data from melanocytes or melanomas. This approach could fail to identify appropriate gene candidates for loci where the causal variant(s) function in a non-melanocytic cell type, e.g. keratinocytes or immune cells for example. Further, cultured melanocytes and melanomas may fail to replicate conditions or cellular contexts under which regulatory elements and causal variants within them may function, and thus may miss interactions with some causal genes. As the degree to which chromatin interactions are stable across such contexts is not entirely clear, interaction data alone may still identify potential candidate genes that only become functional under specific contexts. Lastly, capture-HiC sensitivity and precision are dependent on several factors, including whether a capture bait was designable to any given variant, bait efficiency, the size of the restriction fragment harboring a variant, and bin size used for analyses. Despite considerable apparent sensitivity of this method, some variant to gene interactions could be missed.

## Data Availability

The data produced for this manuscript are being deposited in a public repository and available upon reasonable request.

## ACKNOWLEDGEMENTS

This work has been supported by the Intramural Research Program (IRP) of the Division of Cancer Epidemiology and Genetics, National Cancer Institute, US National Institutes of Health. This work utilized the Biowulf cluster computing system at the NIH. The results appearing here are in part based on data generated by the TCGA Research Network. We would like to thank members at the National Cancer Institute Cancer Genomics Research Laboratory (CGR) for help with sequencing efforts. We also thank all the cohorts, funders, and investigators who contributed to the melanoma GWAS, as originally acknowledged by Landi and colleagues. We would like to thank the research participants and employees of 23andMe. The content of this publication does not necessarily reflect the views or policies of the US Department of Health and Human Services, nor does the mention of trade names, commercial products, or organizations imply endorsement by the US Government. Mark Iles is supported in part by the National Institute for Health and Care Research (NIHR) Leeds Biomedical Research Centre. The views expressed are those of the author(s) and not necessarily those of the NHS, the NIHR or the Department of Health and Social Care.

## AUTHOR CONTRIBUTIONS

R.T., M.X, H.S., J.Yon, L.J., T.M, T.Z., R.C., E.L., T.R., R.H., K.F., J.Yin, M.E.J., A.D.W., A.C., S.F.A.G, M.M.I., M.T.L., M.H.L., J.C., and K.M.B. contributed to the research activities described in this manuscript. K.M.B. led and supervised the research described. M.M. provided additional supervision and mentorship. R.T., M.X., and K.M.B. wrote the manuscript. J. Yon, R.H., M.M., M.M.I., M.H.L., and J.C. provided significant contributions to writing, review, and editing.

## WEB RESOURCES

- CHiCAGO: https://www.functionalgenecontrol.group/chicago
- HiCUP: https://www.bioinformatics.babraham.ac.uk/projects/hicup/
- LDlink: https://ldlink.nih.gov/?tab=home
- Ensembl variant Effect Predictor: https://useast.ensembl.org/info/docs/tools/vep/index.html
- ATAC-seq pipeline: https://github.com/ENCODE-DCC/atac-seq-pipeline
- UCSC browser: https://genome.ucsc.edu
- WashU Epigenome Browser: http://epigenomegateway.wustl.edu
- DAP-G: https://github.com/xqwen/dap
- Bedtools: https://bedtools.readthedocs.io/en/latest/
- Bedtoolsr: http://phanstiel-lab.med.unc.edu/bedtoolsr.html
- ROADMAP epigenomic project: https://egg2.wustl.edu/roadmap/web_portal/chr_state_learning.html
- Intogen: https://www.intogen.org/search
- NIH Biowulf Cluster, http://hpc.nih.gov
- PLINK, https://www.cog-genomics.org/plink/
- The Cancer Genome Atlas (TCGA) Research Network, http://cancergenome.nih.gov/
- TWAS FUSION, http://gusevlab.org/projects/fusion/

## DATA AND CODE AVAILABILITY

Capture-HiC data are being deposited in ArrayExpress, including both called interactions as well as raw sequencing data.

Melanocyte ATAC-seq data are being deposited in ArrayExpress, including called peaks and raw sequencing data.

Data from the 2020 melanoma GWAS meta-analysis performed by Landi and colleagues^10^ were obtained from dbGaP (dbGaP: phs001868.v1.p1), with the exclusion of self-reported data from 23andMe, Inc. and UK Biobank. The full GWAS summary statistics for the 23andMe discovery dataset will be made available through 23andMe to qualified researchers under an agreement with 23andMe that protects the privacy of the 23andMe participants. Please visit https://research.23andme.com/collaborate/#dataset-access/ for more information and to apply to access the data. Summary data from the remaining self-reported cases are available from the corresponding authors of that manuscript (Matthew Law; Mark Iles; and Maria Teresa Landi).

Melanocyte genotype data, RNA-seq expression data, and all eQTL/meQTL association results^23,24^ are accessible through Genotypes and Phenotypes (dbGaP) under accession dbGaP: phs001500.v2.p1. The MPRA sequencing^25^ and associated MPRA sequencing data are accessible through Gene Expression Omnibus (GEO; https://www.ncbi.nlm.nih.gov/geo/) under the accession GEO: <u>GSE210356</u>.

