## Supplementary Figures for "Mapping chromatin interactions at melanoma susceptibility loci and cell-type specific dataset integration uncovers distant gene targets of *cis*-regulation"

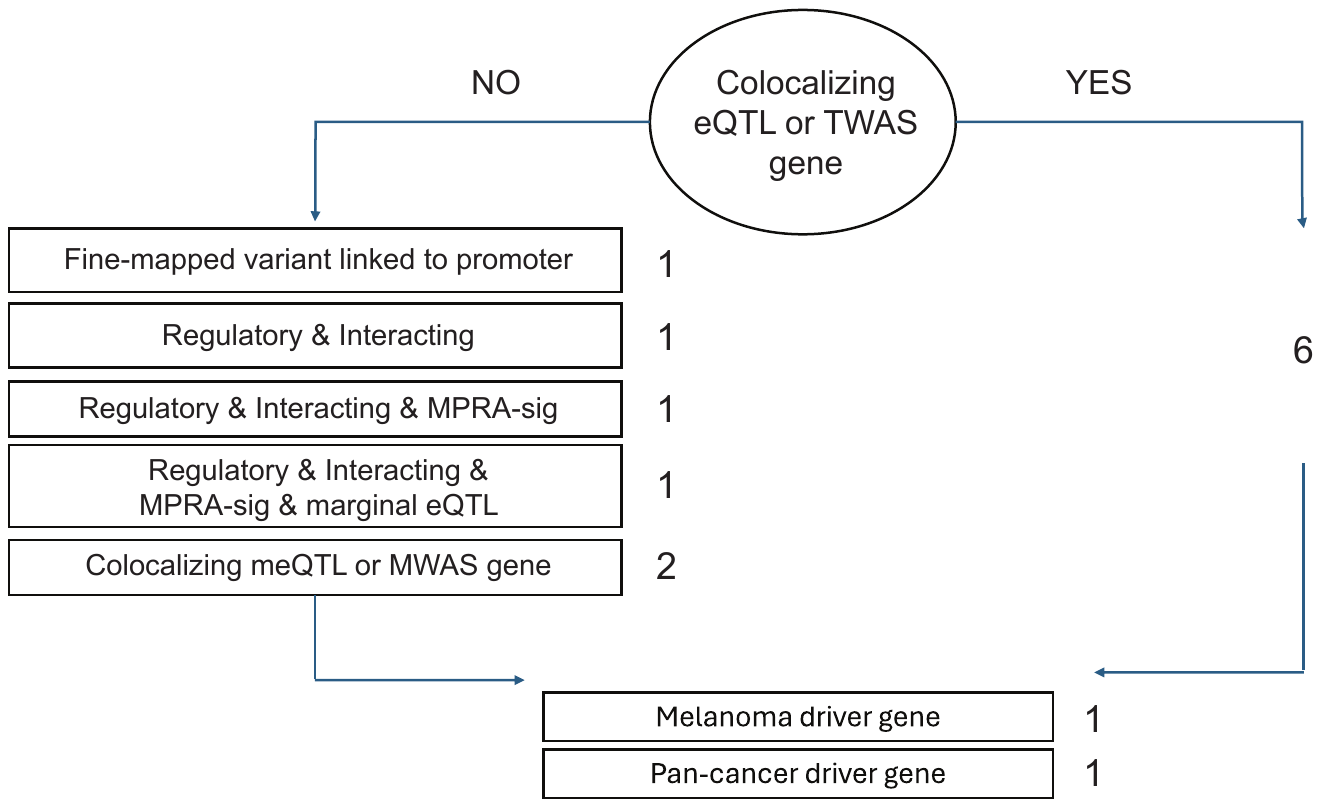
­­_­_

**Supplementary Figure 1. Integrative scoring schematic for nominated candidate genes at melanoma GWAS risk signals**. We strongly weighted prior evidence of a colocalizing eQTL or an FDR-significant TWAS finding for a gene, assigning such genes a score of 6. For genes without such QTL data, we cumulatively scored potential candidate genes up to a score of 6 based on a fine-mapped variant interacting with or being physically located within a gene promoter (+1), the interacting fine-mapped variant being located within an annotated melanocyte or melanoma regulatory region (+1), the interacting fine-mapped variant showing a significant allelic cis-regulatory effect in massively parallel reporter assays (+1), as well as whether a CpG probe within a gene promoter or body showed a colocalizing meQTL or FDR-significant MWAS result (+2). For all genes, we further scored them if they were previously identified as a melanoma (+1) or pan-cancer driver gene (+1).

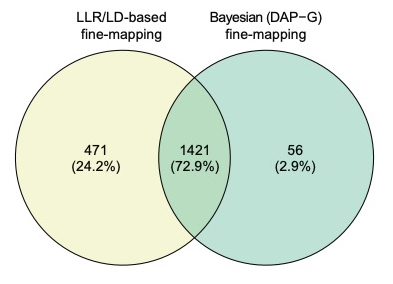

**Supplementary Figure 2. Number of fine-mapped credible causal variants (CCVs) across all 68 melanoma GWAS signals using Bayesian (DAP-G) or log-likelihood ratio (LLR)/LD-based fine-mapping methods**. The Venn diagram shows the overlap between fine-mapped variants identified by the LLR/LD approaches and the Bayesian DAP-G approach.

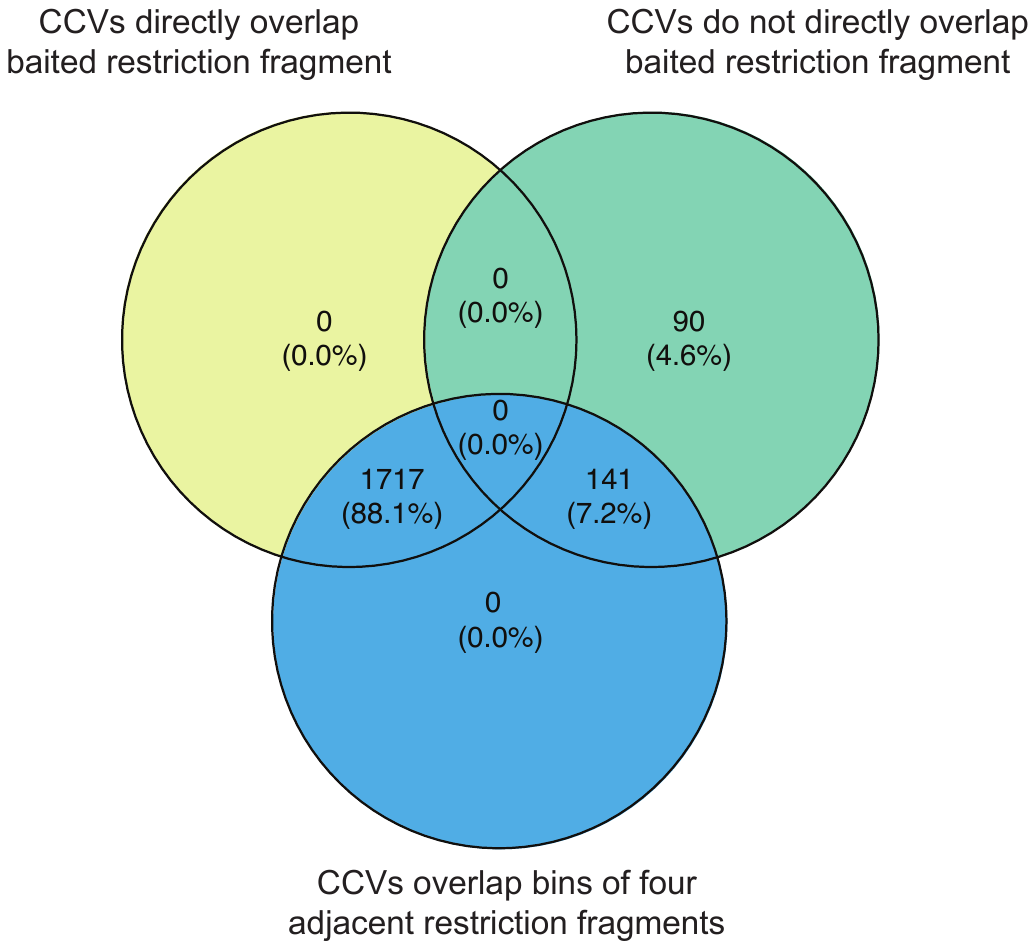

**Supplementary Figure 3. Venn diagram showing number of fine-mapped CCVs that directly overlap a successfully baited restriction fragment in the custom capture-HiC assay design**. 1,717 out of the total 1948 fine-mapped variants directly overlapped a baited restriction fragment and were assessable in the one-fragment (1F) analysis. For 4F analysis, there was slightly better coverage of fine-mapped CCVs, as 1858 CCVs were located within four fragment bins that were directly baited.

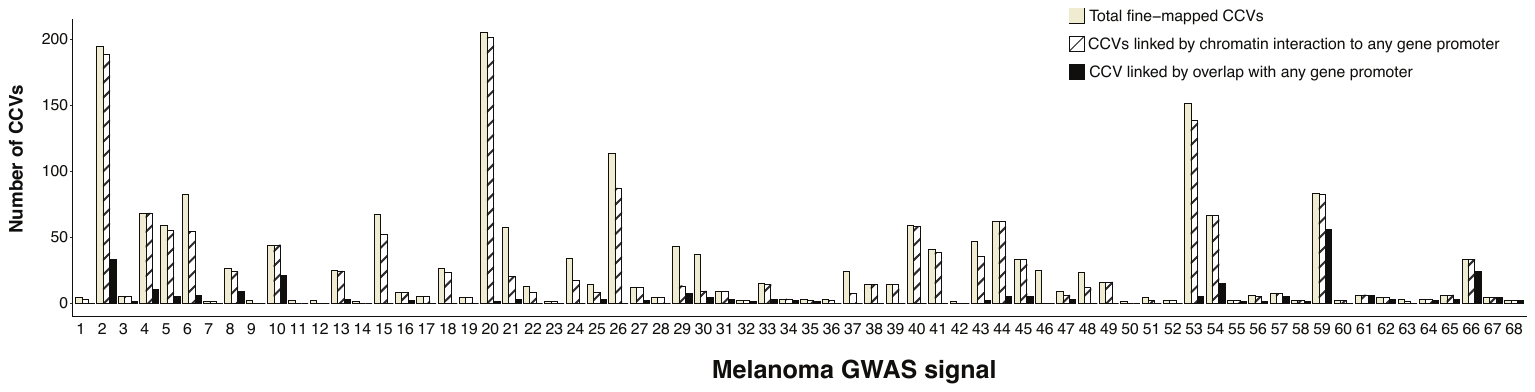

**Supplementary Figure 4. Summary of fine-mapped variant physically interaction with or located within in at least one gene promoter**. The ‘All fine-mapped CCVs’ represents the total number of fine-mapped variants at each of the 68 signals. The ‘CCV with chromatin interaction to promoter’ set represents the number of fine-mapped variants found to interact with at least one gene promoter, whether melanocyte- or melanoma-specific, or a more globally defined set. The ‘CCV located in promoter’ set represents the number of fine-mapped variants directly located within at least one gene promoter region the annotated global, or melanocyte-specific, or melanoma-specific gene promoter(s).

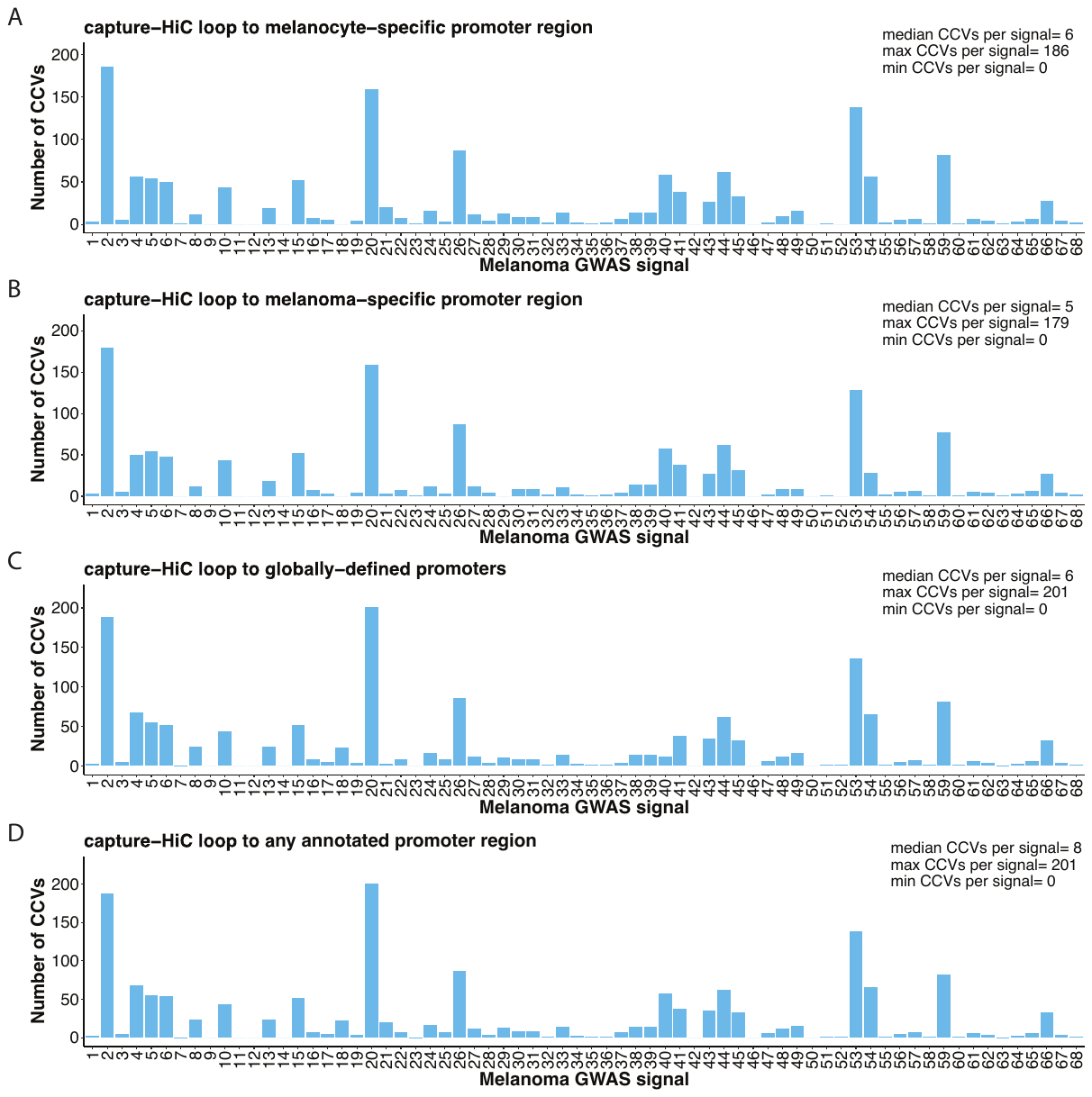

**Supplementary Figure 5. Summary of the number of fine-mapped credible causal variants (CCVs) with detected capture-HiC chromatin interaction loops**. Shown are the number of CCVs with interaction to at least one (A) melanocyte-specific promoter region, (B) melanoma-specific promoter region, (C) globally defined promoter region, or (D) any promoter region across the 68 independent melanoma GWAS risk signals

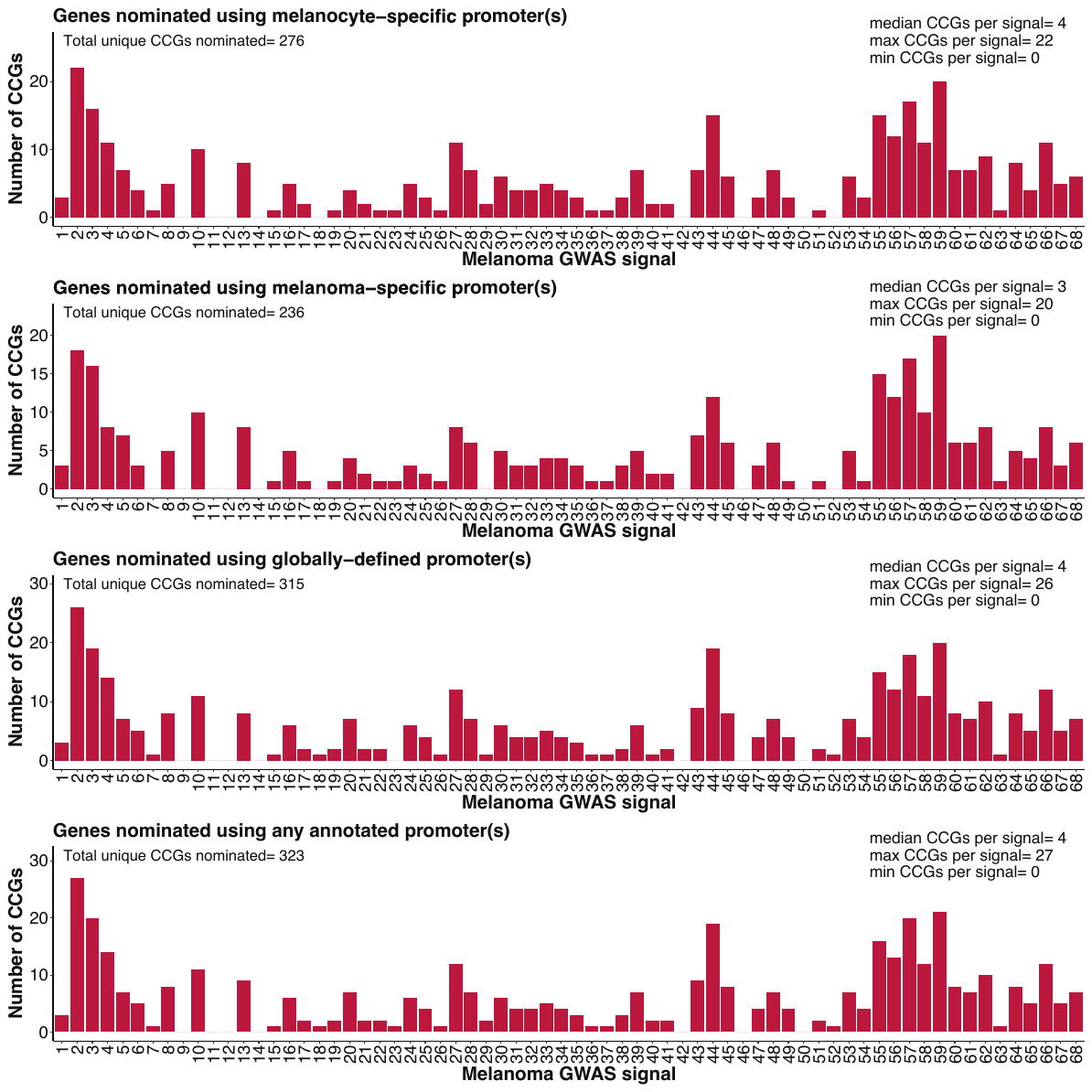

**Supplementary Figure 6. Summary of the number of candidate causal genes (CCGs) nominated via chromatin interaction**. Shown are the number of CCGs nominated via interaction with melanocyte-specific promoter region(s), melanoma-specific promoter region(s), globally-defined promoter region(s), or any annotated promoter region(s) across the 68 independent melanoma GWAS risk signals. Minimum, maximum, and median number of genes per risk signal are shown.

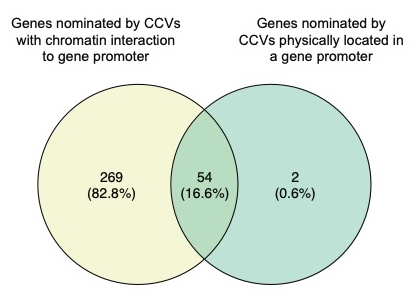

**Supplementary Figure 7. Overlap between genes nominated by chromatin interaction and those nominated by CCVs physically located within a promoter across all melanoma GWAS signals**.

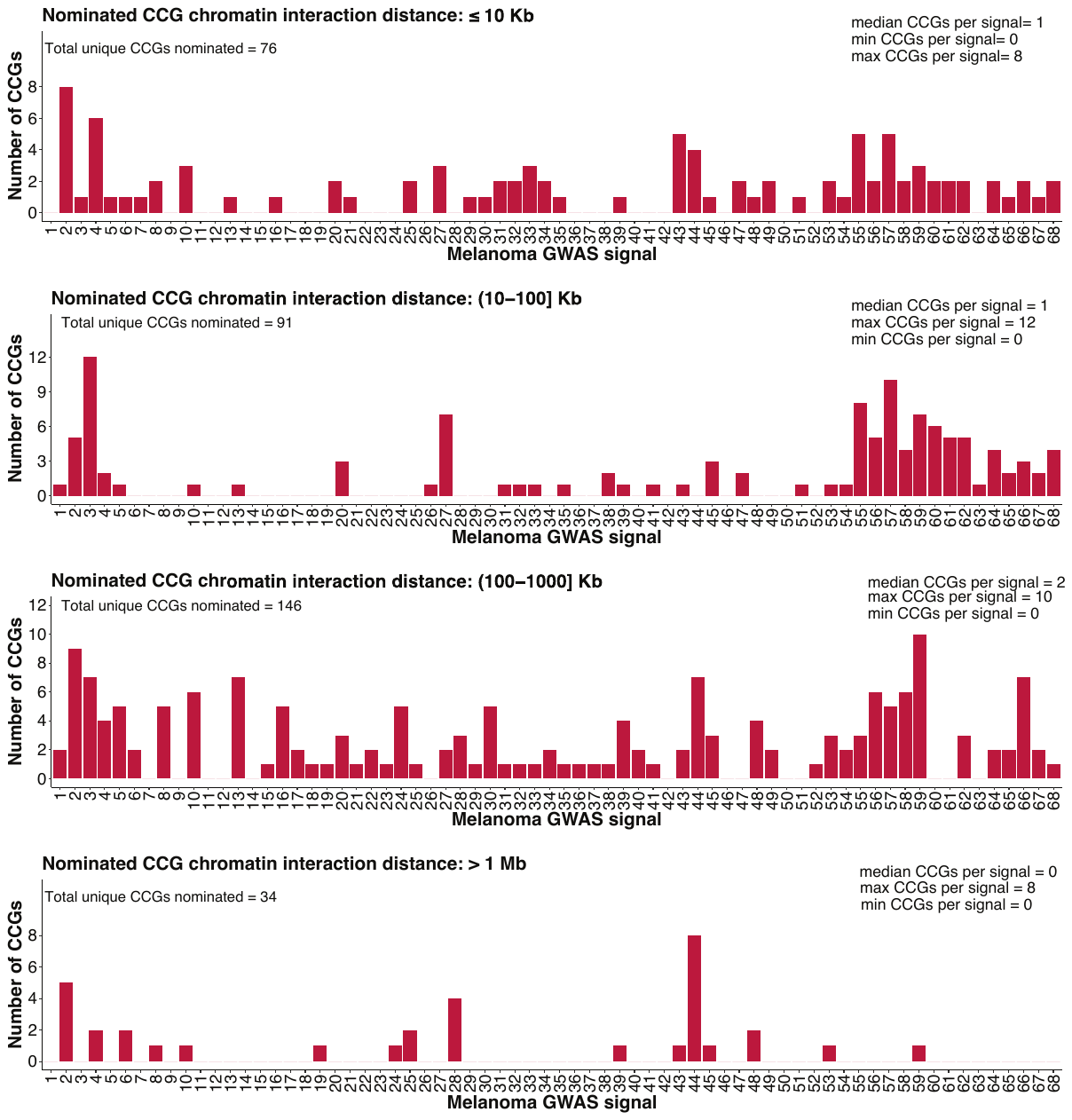

**Supplementary Figure 8. Bar plot summarizing the number of candidate genes nominated by chromatin interaction of varying distance across 68 GWAS signals.**

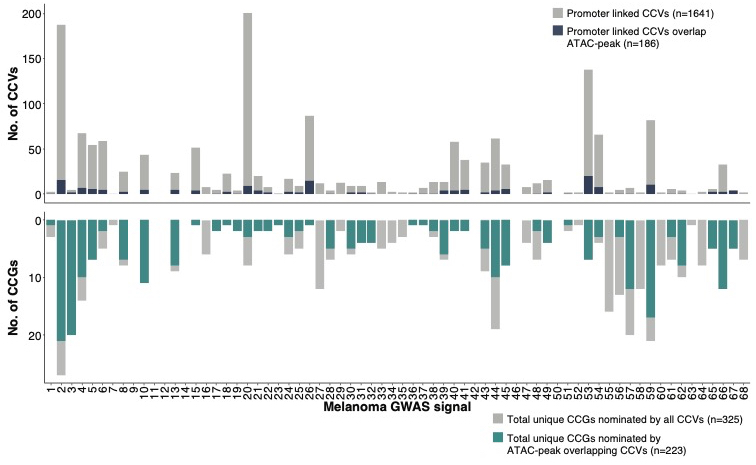

**Supplementary Figure 9: Stacked bar plot summary of fine-mapped variants (CCVs) and nominated target genes (CCGs) after integrating the chromatin interaction dataset with melanocyte- and melanoma-specific ATAC-seq datasets for each of 68 melanoma risk signals**. The top bar plot shows the total number of fine-mapped variants that are linked to at least one target gene using the chromatin interaction dataset, while blue color shows the number of interacting variants overlapping open chromatin in any of the ATAC-seq datasets. The bottom plot shows the number unique genes nominated as potential candidates using chromatin interaction data only, while the green shows the number of candidate genes following integration with ATAC-seq data.

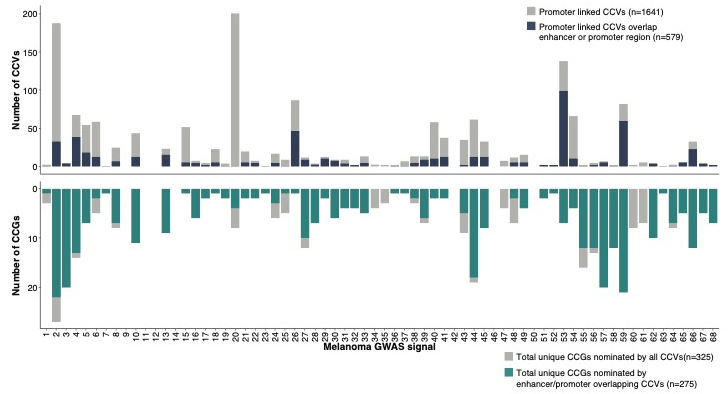

**Supplementary Figure 10: Stacked bar plot summary of fine-mapped variants (CCVs) and nominated target genes (CCGs) after integrating the chromatin interaction dataset with melanocyte- and melanoma-specific enhancer, promoter annotations from chromHMM datasets for each of 68 melanoma risk signals**. The top bar plot shows the total number of fine-mapped variants that are linked to at least one target gene using the chromatin interaction dataset, while blue color shows the number of interacting variants overlapping enhancer or promoter in any of the chromHMM datasets. The bottom plot shows the number unique genes nominated as potential candidates using chromatin interaction data only, while the green shows the number of candidate genes nominated following integration with chromHMM dataset.

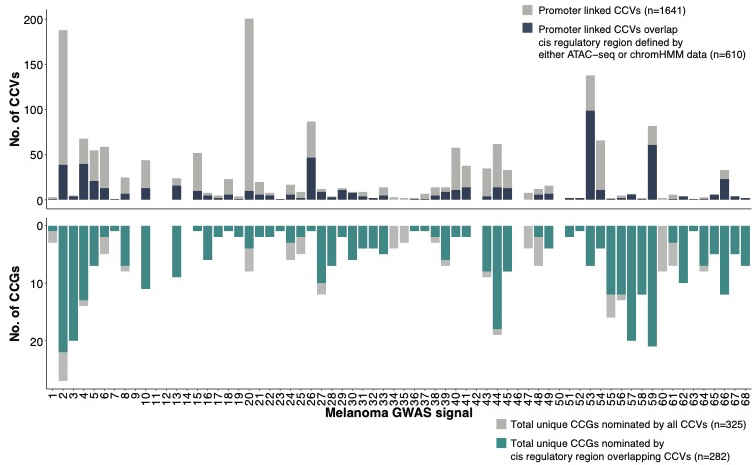

**Supplementary Figure 11: Stacked bar plot summary of fine-mapped variants (CCVs) and nominated target genes (CCGs) after integrating the chromatin interaction dataset with melanocyte- and melanoma-specific ATAC-seq and chromHMM datasets for each of 68 melanoma risk signals**. The top bar plot shows the total number of fine-mapped variants that are linked to at least one target gene using the chromatin interaction dataset, while blue color shows the number of interacting variants overlapping a potential regulatory region in any of the ATAC-seq or chromHMM datasets. The bottom plot shows the number unique genes nominated as potential candidates using chromatin interaction data only, while the green shows the number of candidate genes nominated following integration with ATAC-seq and chromHMM datasets.

**Supplementary Figure 12: Integrative evidence for candidate causal genes at melanoma risk signals.** (A-H) For each locus, the figure indicates the nearest gene to the lead variant, summarizes candidate gene expression in primary melanocytes and melanoma tumors, indicates genes implicated by interaction of fine-mapped variants to the gene’s promoter, along with further refined evidence for these interacting variants integrated with melanocyte and melanoma epigenomic and MPRA data. Also summarized are melanocyte eQTL/TWAS evidence, meQTL/MWAS evidence, and whether the candidate gene has been implicated as a melanoma or pan-cancer driver gene. Finally, the figures show an overall integrative score for each candidate scored from 0-8 with 8 being the highest score.

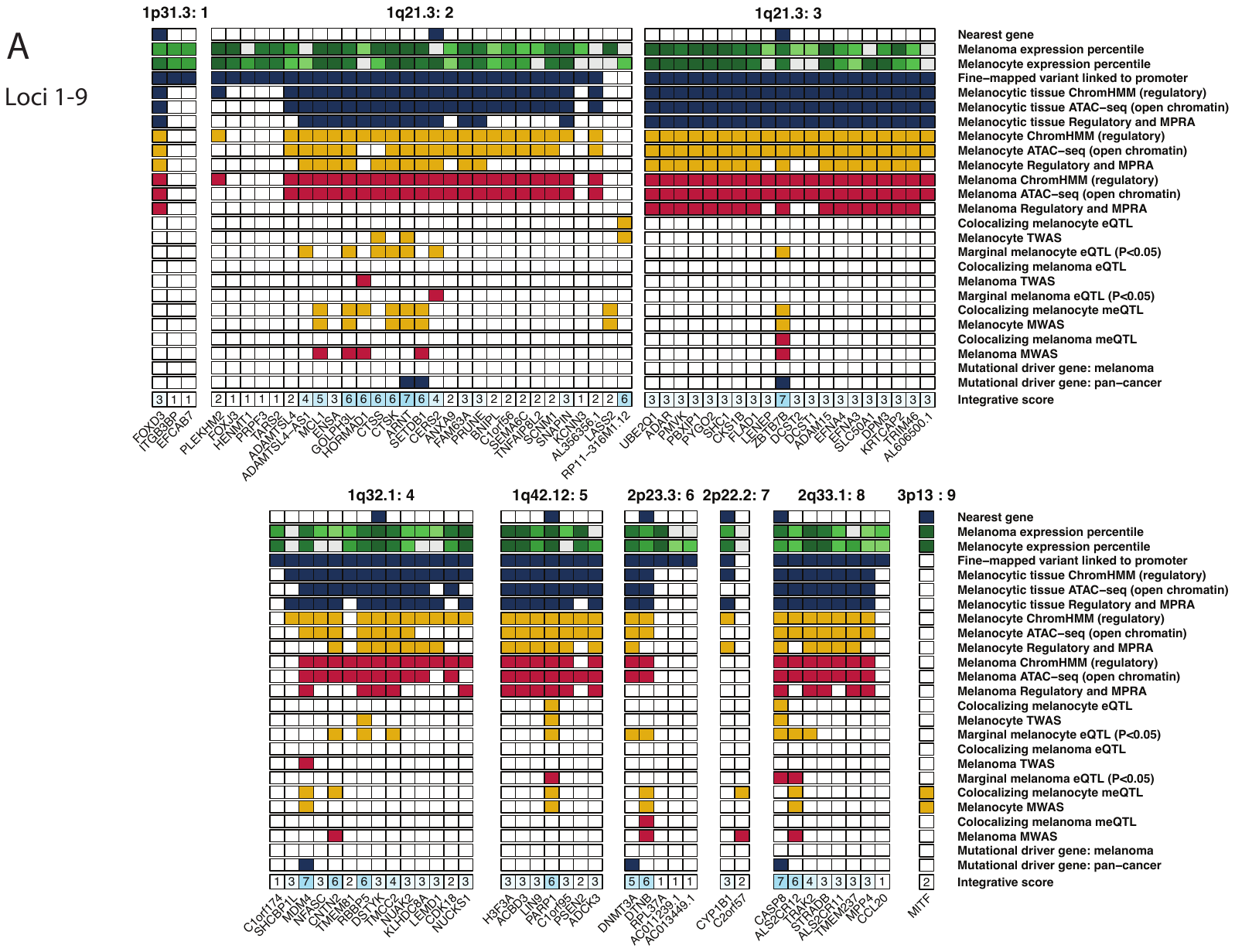

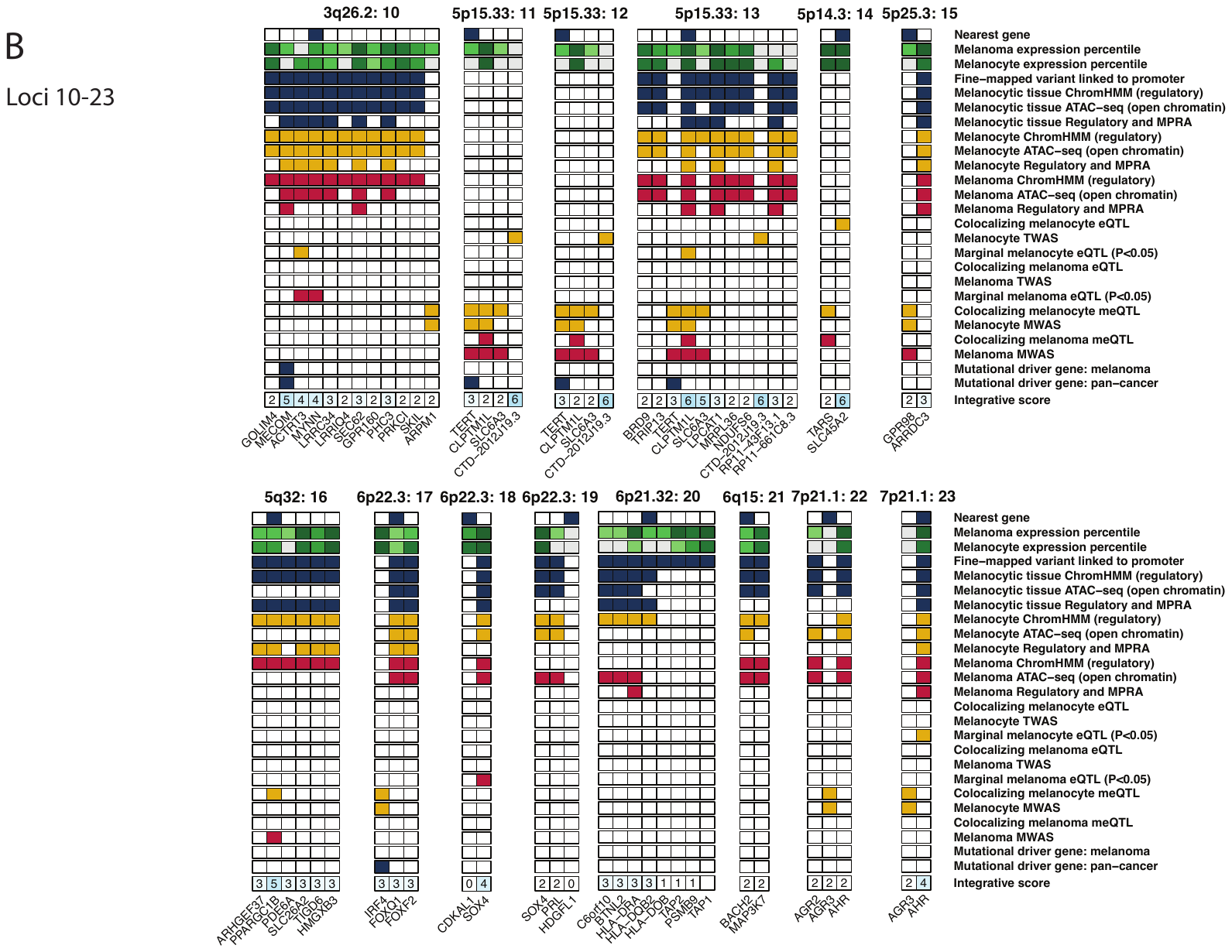

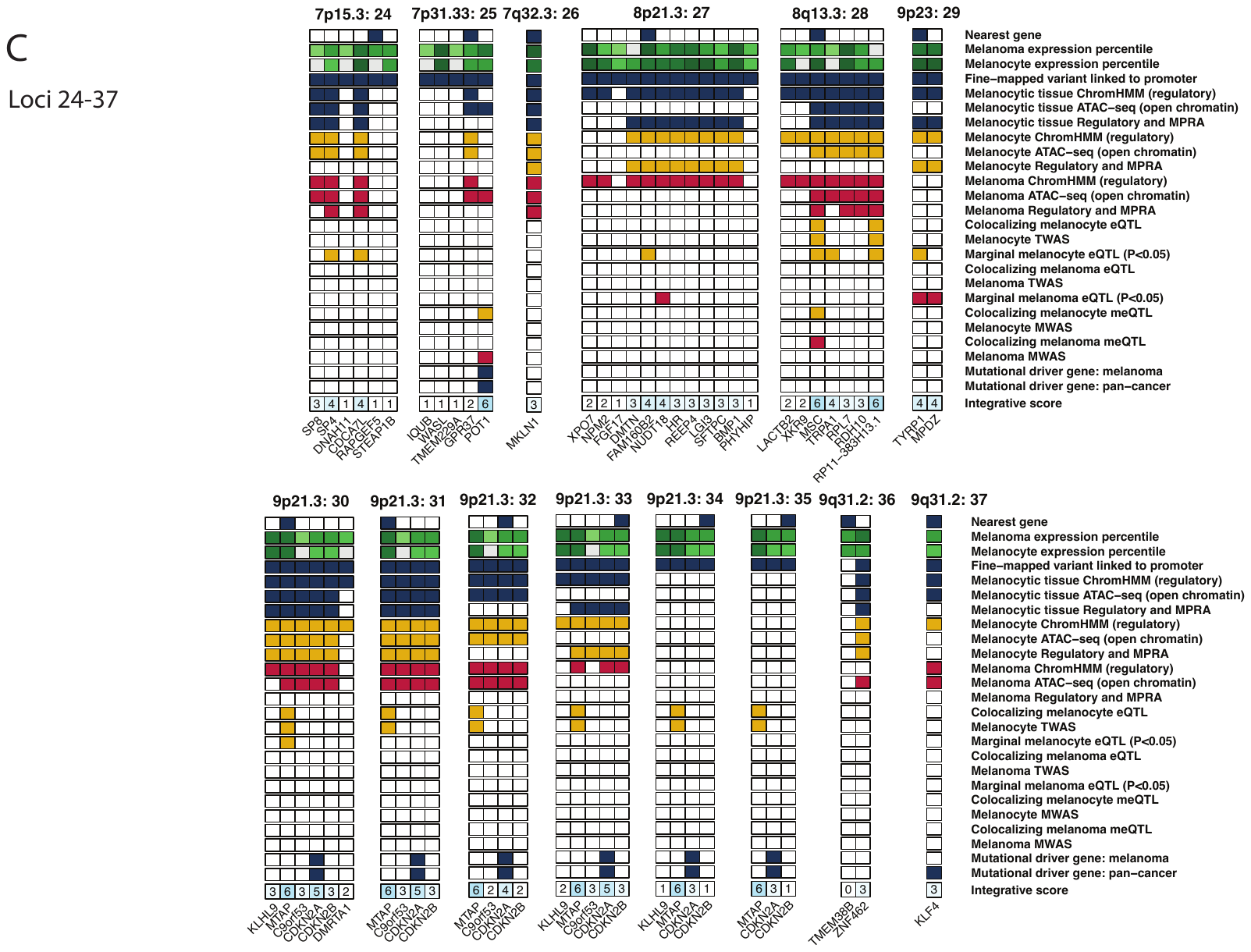

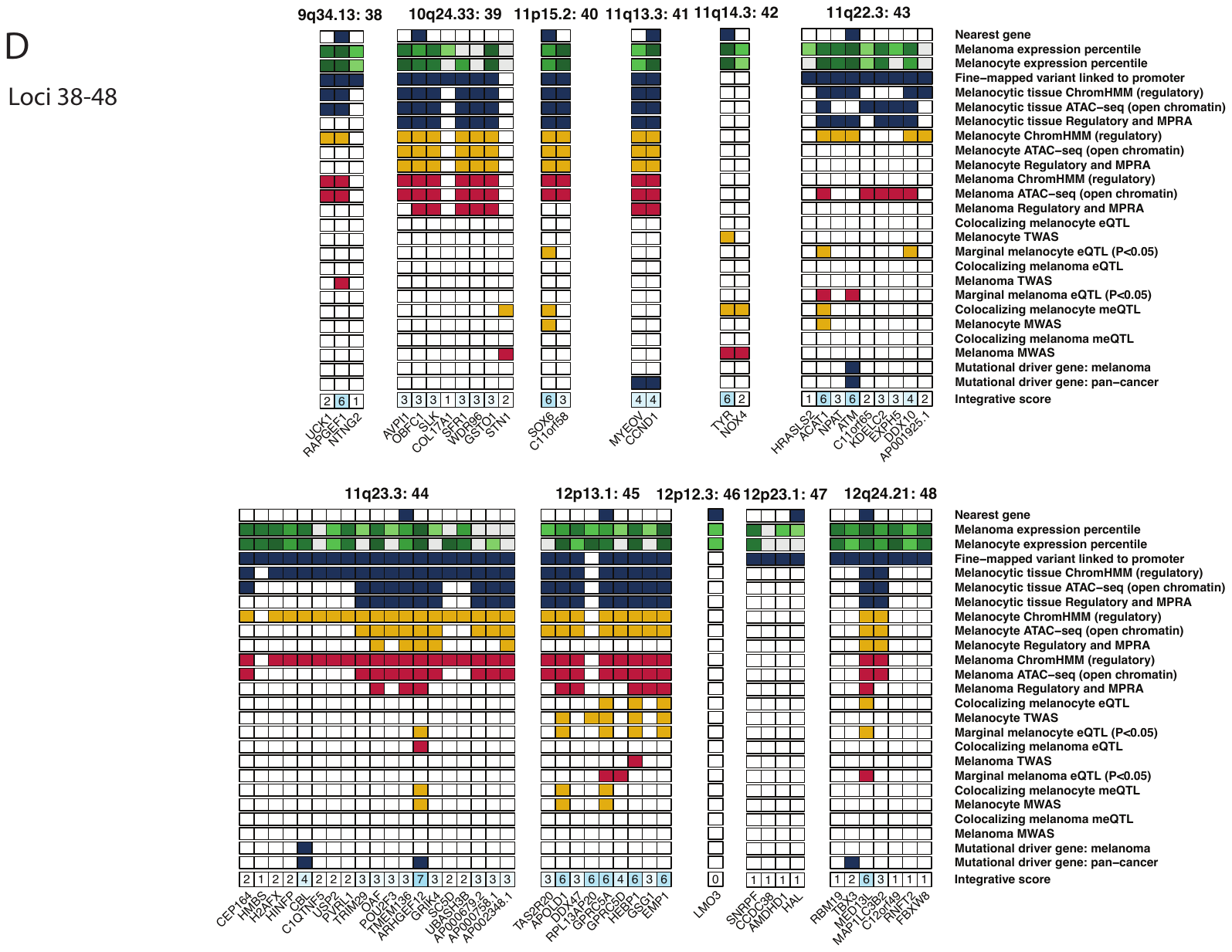

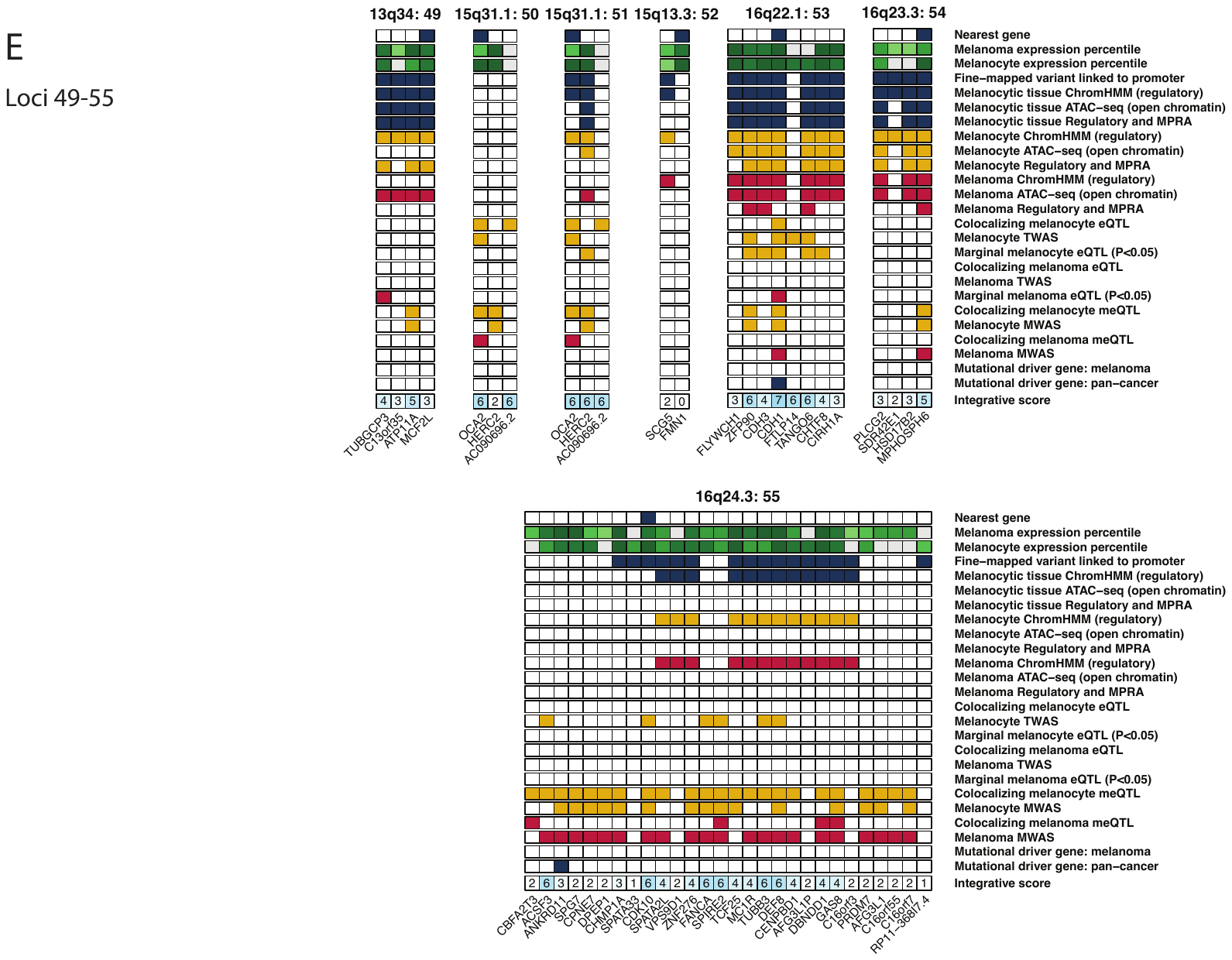

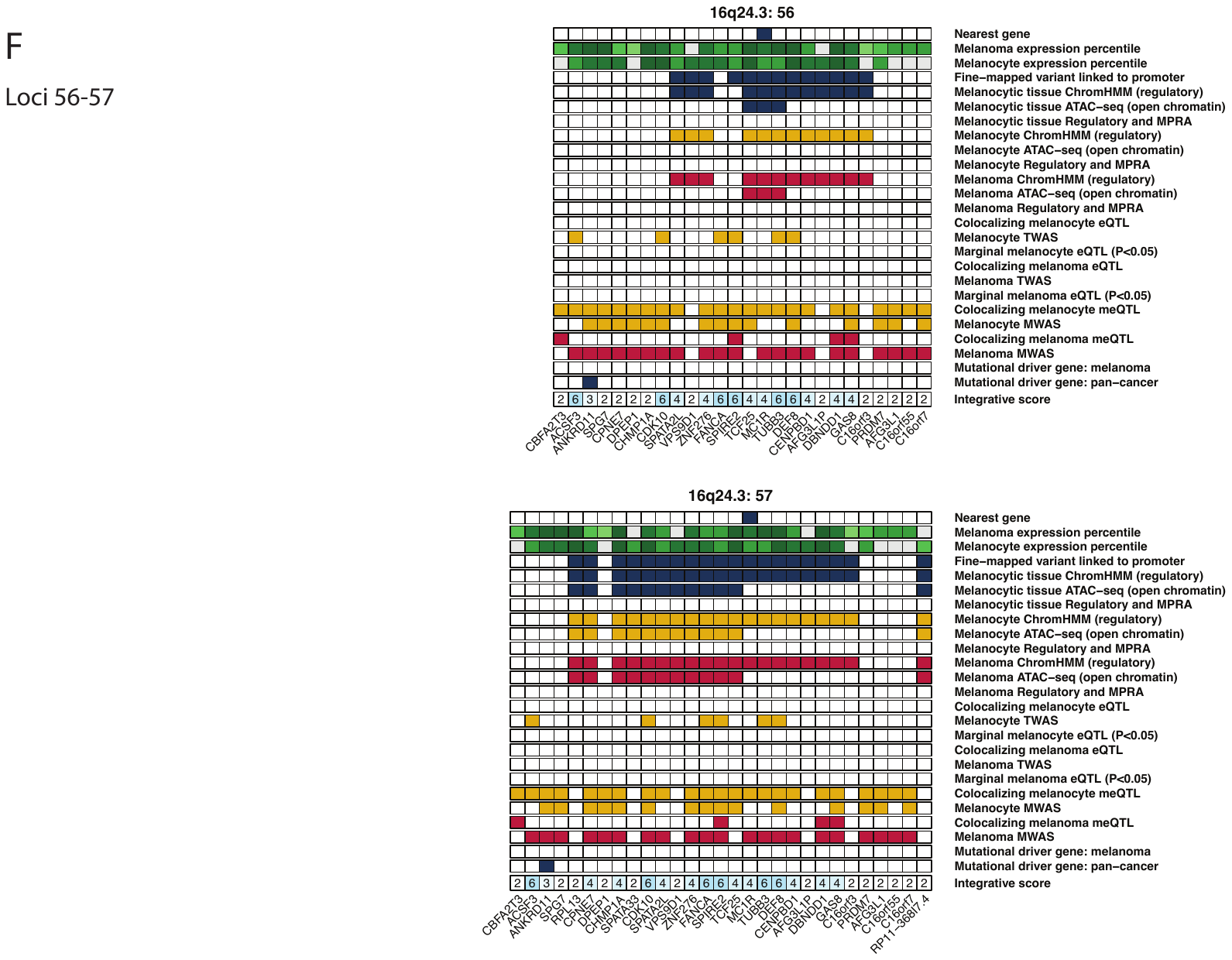

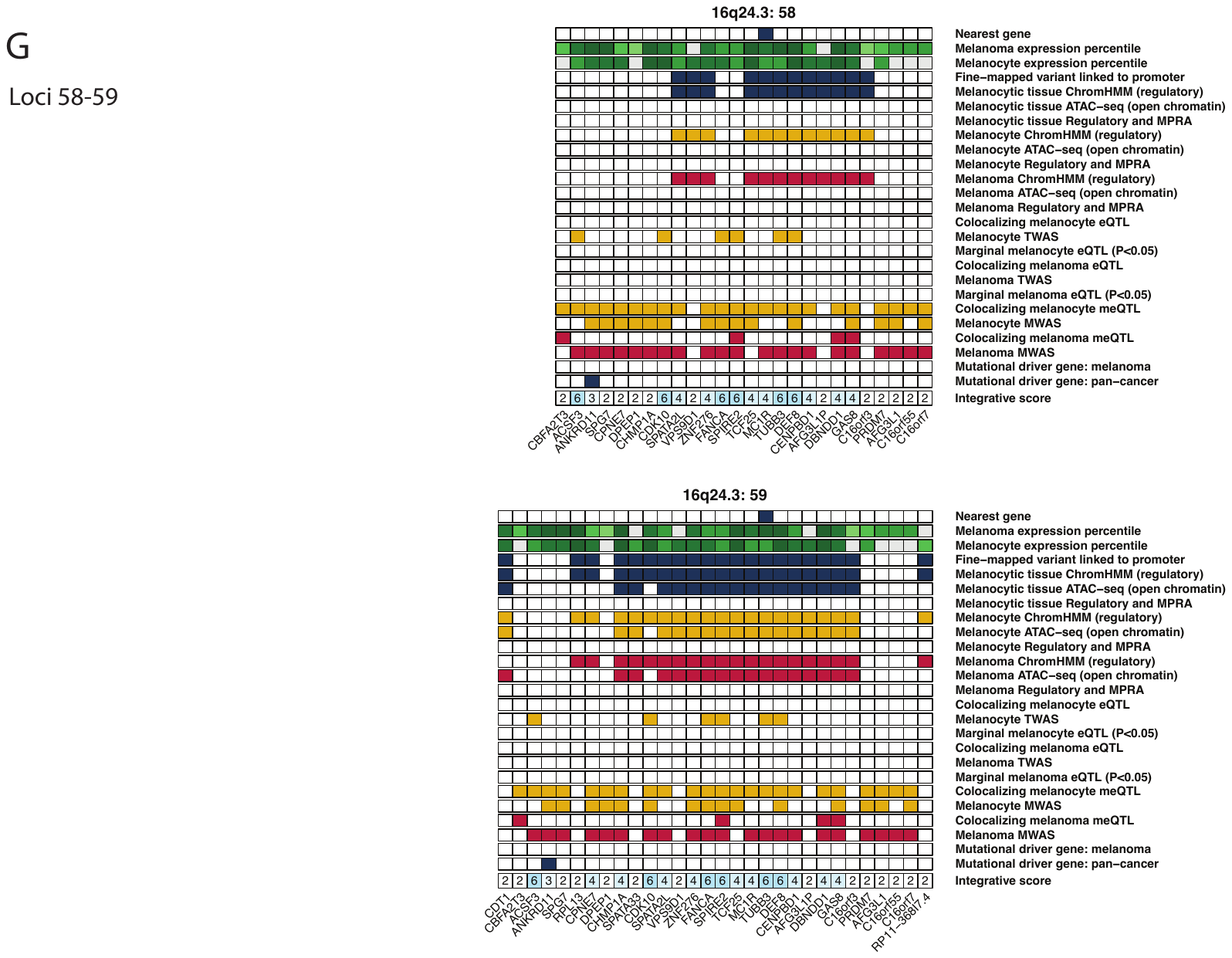

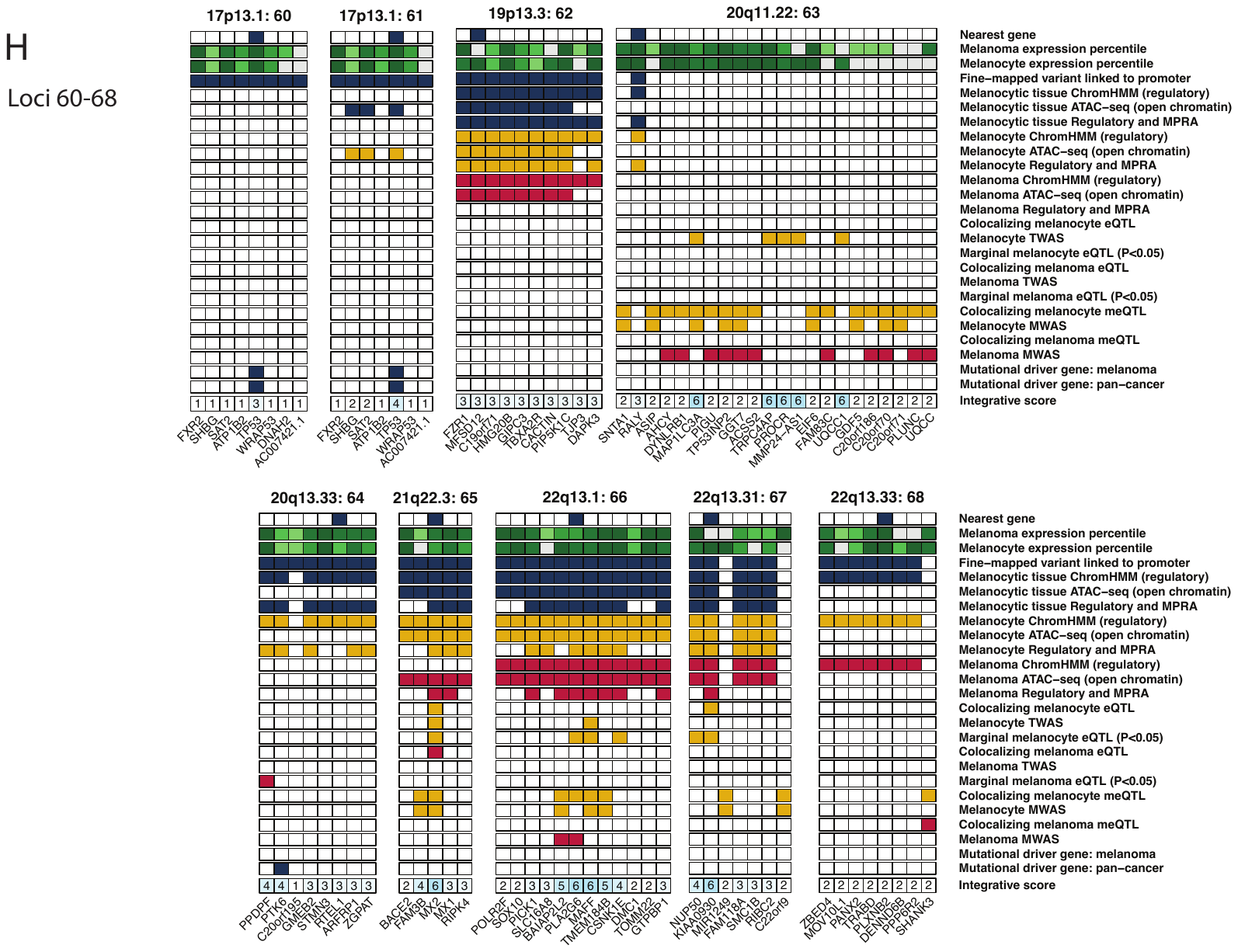

**Supplementary Figure 13: Chromatin looping from loci (A) overlapping SOX6 on chromosome 11, (B) overlapping RBBP5 and DSTYK on chromosome 1, and (C) overlapping ARHGEF12 and chromosome 11.** Figure shows data from melanocyte DNAseI hypersensitivity sequencing (Roadmap, n=2 melanocyte cultures), melanocyte ChromHMM (Roadmap, n=2 melanocyte cultures), melanocyte ATAC-seq (n=5 cultures), and melanoma cell ATAC-seq relative to genes in the region. Fine-mapped variants for both loci and location of capture-HiC baits is shown along with chromatin looping. Potentially regulatory fine-mapped variants from (A) the locus overlapping SOX6 shows interactions with the SOX6 promoter. Potentially regulatory fine-mapped variants from (B) the locus overlapping RBBP5 and DSTYK physically interact with the promoter of MDM4. Finally, potentially regulatory fine-mapped variants at the locus overlapping ARHGEF12 interact with the promoter of CBL.

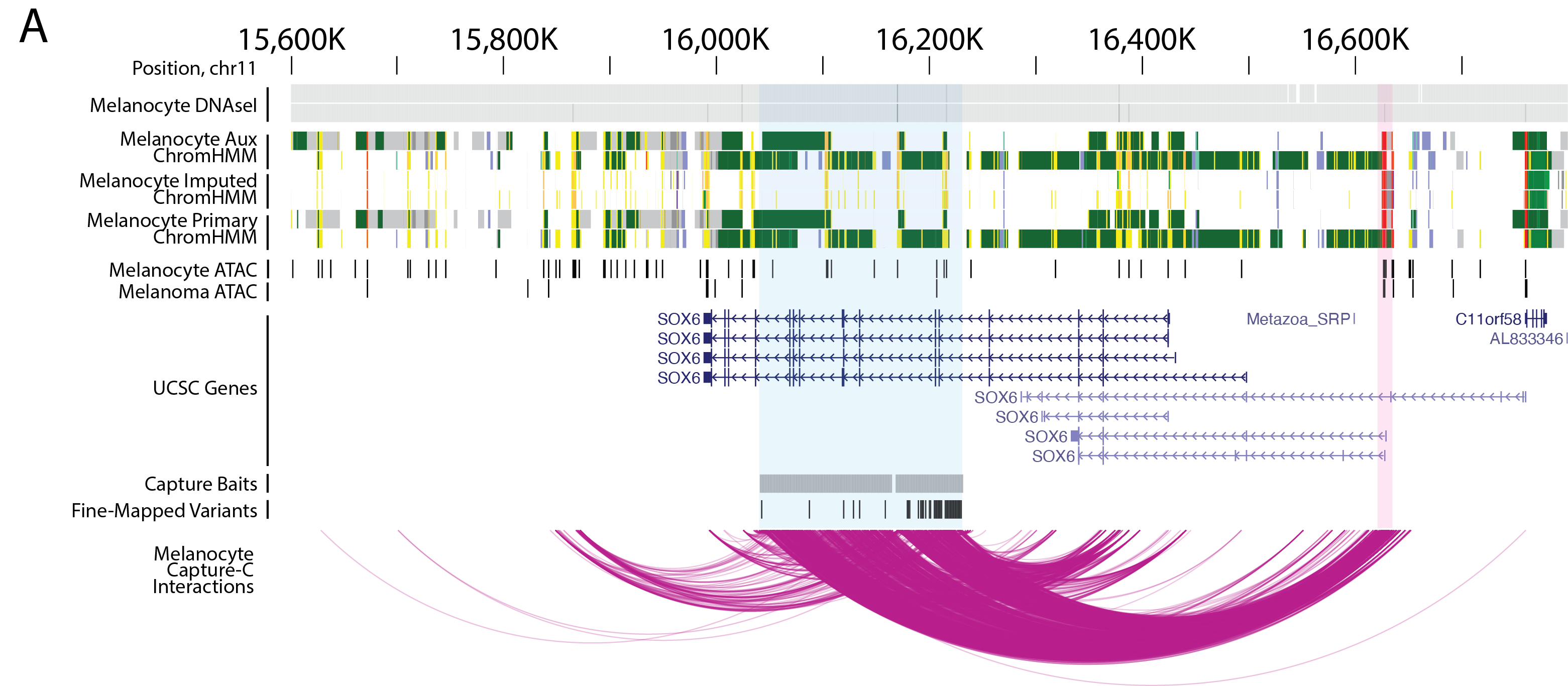

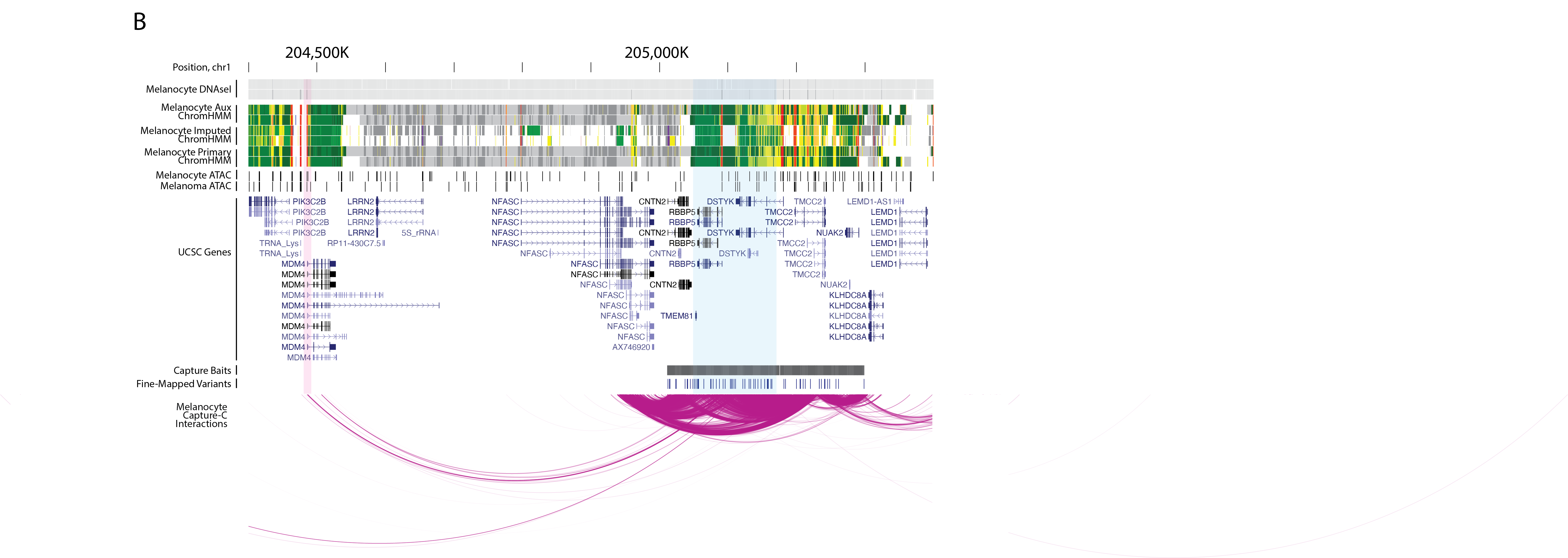

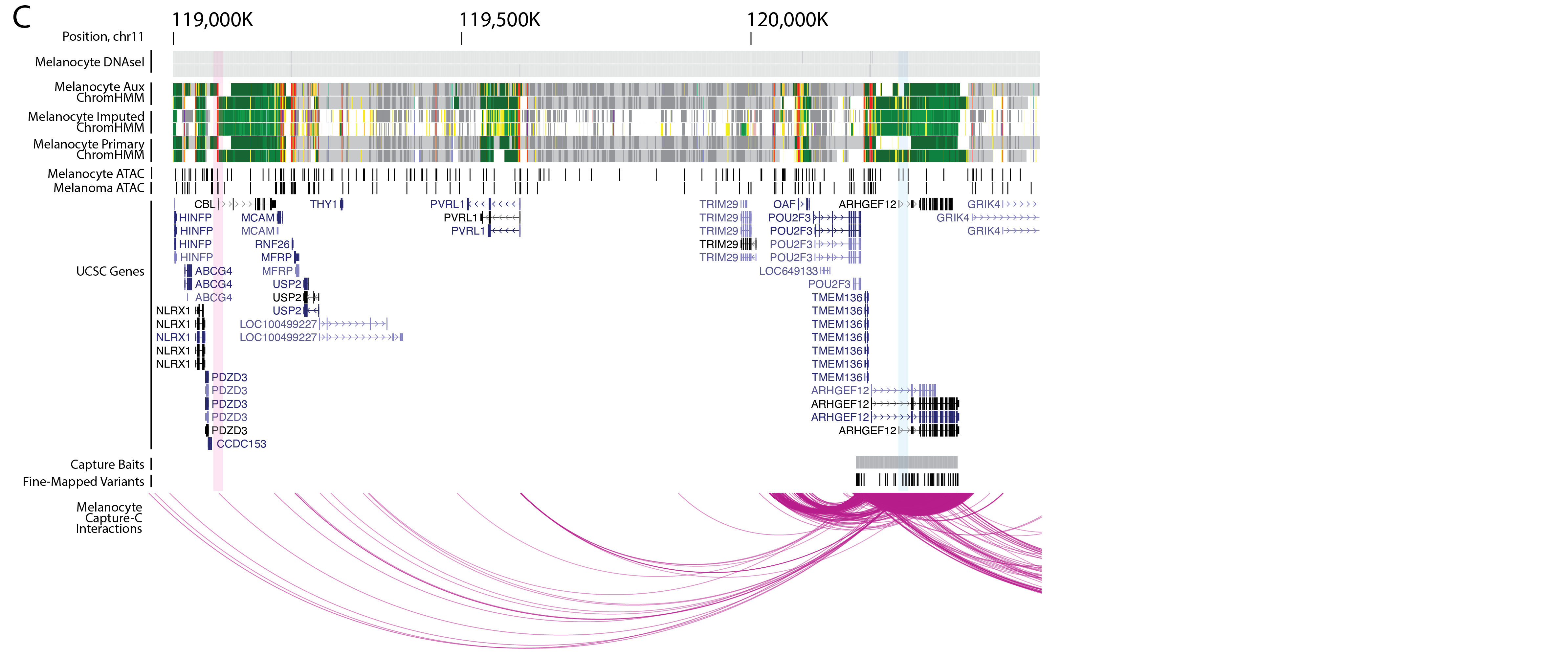

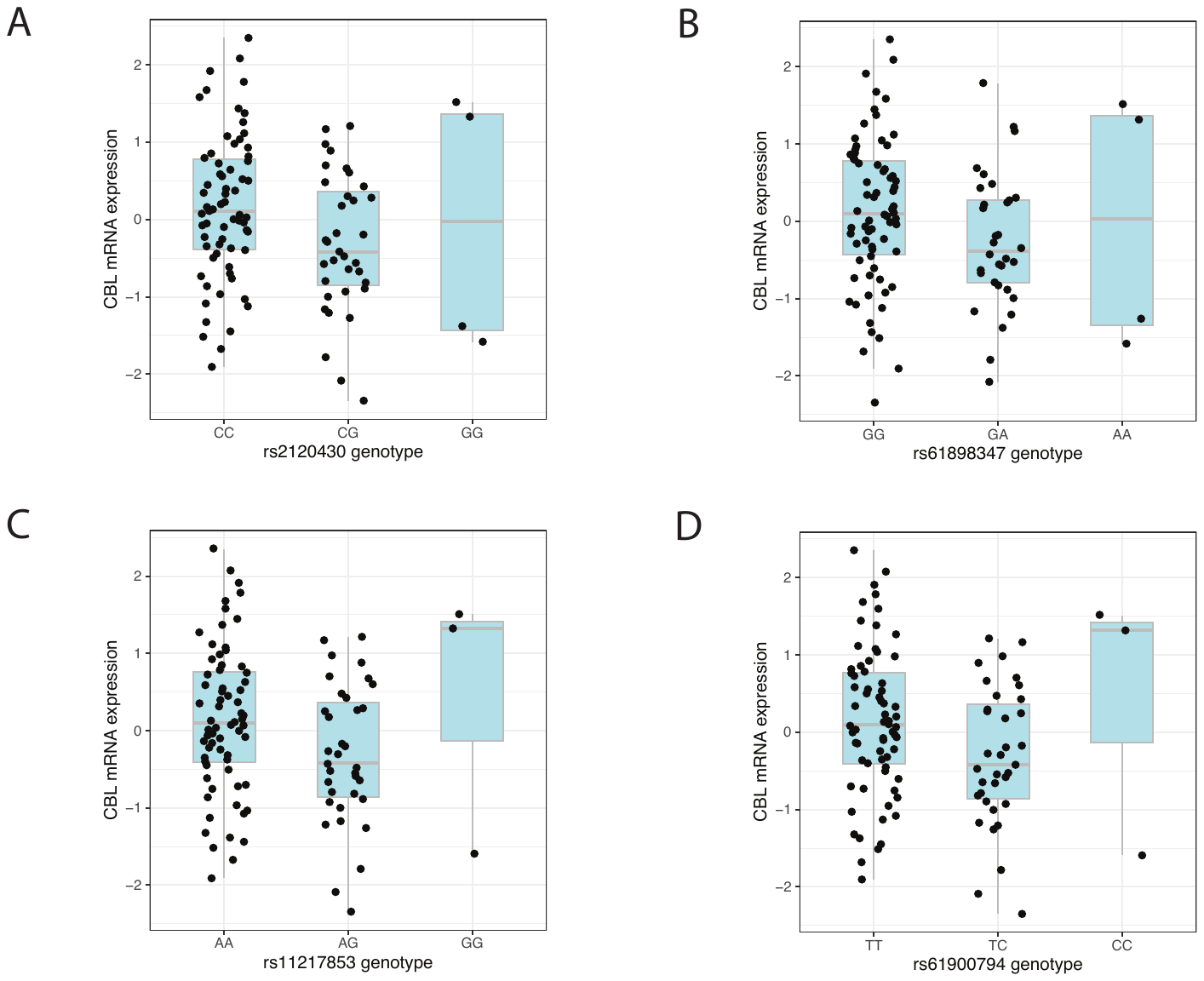

**Supplementary Figure 14. eQTL analysis for candidate causal sequence variants for locus 36 (signal 44) on chromosome 11 and CBL in melanocytes.** Plots showing CBL expression levels in 106 primary human melanocyte cultures grouped by genotype of (A) rs2120430, (B) rs61898347, (C) rs11217853, and (D) rs61900794. Homozygotes for the risk allele are shown to the right, while homozygotes for the protective alleles to the left of each graph.
